# Cardiovascular risk prediction - a systems medicine approach

**DOI:** 10.1101/2023.03.16.23287363

**Authors:** Ingrid Gergei, Thomas Pfau, Bernhard K. Krämer, Jochen G. Schneider, Thanh Phuong Nguyen, Winfried März, Thomas Sauter

## Abstract

**Background:** Guidelines for the prevention of cardiovascular disease (CVD) have recommended the assessment of the total CVD risk by risk scores. Current risk algorithms are low in sensitivity and specificity and they have not incorporated emerging risk markers for CVD. We suggest that CVD risk assessment can be still improved. We have developed a long-term risk prediction model of cardiovascular mortality in patients with stable coronary artery disease (CAD) based on newly available machine learning and on an extended dataset of new biomarkers.

**Methods:** 2953 participants of the Ludwigshafen Risk and Cardiovascular Health (LURIC) study were included. 184 laboratory and 21 demographic markers were ranked according to their contribution to risk of cardiovascular (CV) mortality using different data mining approaches. A self-learning bioinformatics workflow, including seven different machine learning algorithms, was developed for CV risk prediction. The study population was stratified into patients with and without significant CAD. Thereby, significant CAD was defined as a lumen narrowing of 50 % or more in at least one of the coronary segments or a history of definite myocardial infarction. The machine learning models in both subpopulations were compared with established CV risk assessment tools.

**Results:** After a follow-up of 10 years, 603 (20.4%) patients died of cardiovascular causes. 95 (%) patients without CAD deceased within ten years and 247 (13.2 %) patients with CAD within 5 years. Overall and in patients without CAD, NT-proBNP (N-terminal pro B-type natriuretic peptide), TnT (Troponin T), estimated cystatin c based GFR (glomerular filtration rate) and age were the highest ranked predictors, while in patients with CAD, NT-proBNP, GFR, CT-proAVP (C-terminal pro arginine vasopressin) and TNT were highest predictive. In the comparison with the FRS, PROCAM and ESC risk scores, the machine learning workflow produced more accurate and robust CV mortality prediction in patients without CAD. Equivalent CV risk prediction was obtained in the CAD subpopulation in comparison with the Marschner risk score. Overall, the existing algorithms in general tend to assign more patients into the medium risk groups, while the machine learning algorithms tend to have a clearer risk/no risk assignment. The framework is available upon request.

**Conclusion:** We have developed a fully automated and self-validating computational framework of machine learning techniques using an extensive database of clinical, routinely and non-routinely measured laboratory data. Our framework predicts long-term CV mortality at least as accurate as existing CVD risk scores. A combination of four highly ranked biomarkers and the random forest approach showed the best predictive results. Moreover, a dynamic computational model has several advantages over static CVD risk prediction tools: it is freeware, transparent, variable, transferable and expandable to any population, types of events and time frames.

## Introduction

Cardiovascular disease (CVD) is still the leading cause of death (1). Worldwide, 17.6 million people died from cardiovascular disease in 2016 of which ischemic heart disease and stroke together accounted for 85.1% of all deaths (1). Studies in the past have shown that at least 50% of CVDs would be possible to prevent by tracking unhealthy lifestyle and optimizing risk factors (2, 3).

In the past, more than 100 different risk scores have been developed (10). According to US American and European guidelines for cardiovascular disease prevention, the intensity of drug treatment in primary prevention depends on the assessment of an individual’s CV risk (4, 5). For patients with lower CV risk and in a primary prevention setting, the European guideline recommend the use of the SCORE system (6); further risk assessment systems, including the Framingham Risk Score (FRS) (7) and the PROCAM Risk Score (8) are available. For patients with high to very high CVD risk, the Marschner risk score (9) has been suggested.

Investigations which compared different risk scores have revealed that one of three calculators might classify a patient in a wrong category (11). Moreover, few of the risk calculators were validated sufficiently (10–12).

The diversity and limitations of the available risk equations has prompted us to develop a cardiovascular risk assessment tool based on a novel workflow using machine learning, which calculates each individuaĺs CVD risk rapidly, accurately and in a fully automatized way using laboratory and clinical data. We have performed our analysis in a large cohort of patients initially presenting for coronary angiography. We hypothesized that a bioinformatics approach might enhance cardiovascular risk prediction in comparison to conventional statistical methods and might be easier to implement in laboratory and clinical information systems.

## Materials and Methods

### Study design

We studied participants of the Ludwigshafen Risk and Cardiovascular Health (LURIC). The study protocol has been published (13). In brief, 3316 participants of German ancestry were enrolled between July 1997 and January 2000. Only patients with a coronary angiogram were included. Coronary artery disease (CAD) was assessed by coronary angiography based on maximal luminal narrowing upon visual assessment. All participants were followed over a median observation period of 9.9 years. Written informed consent was obtained from each participant prior to inclusion. The study was in accordance with the Declaration of Helsinki and approved by the ethics committee at the Medical Association of Rheinland-Pfalz (Ärztekammer Rheinland-Pfalz).

### Laboratory test

Blood was drawn in the morning hours at the Heart Centre Ludwigshafen, Germany, immediately centrifuged to obtain EDTA plasma and stored at -80°C for later analysis. A set of 184 metabolic markers were considered in the current analyses. Estimated glomerular filtration rate was determined based on cystatin C levels using the Chronic Kidney Disease Epidemiology Collaboration formula (14). The biomarker methodology and results from the LURIC study have been published previously (13, 15).

### Clinical definitions

A set of 21 clinical markers was used. They included BMI, waist-to-hip ratio, smoking, family history of myocardial infarction, diabetes mellitus defined as HbA1c > 6.5%, clinical measurements such as blood pressure, heart rate, left ventricular ejection fraction determined by echocardiography and the use of commonly used drugs ACE inhibitors, Angiotensin II receptor blockers, β-blockers, calcium channel blockers, diuretics, statins, antidiabetic drugs, platelet inhibitors and gout-treatment.

### Exclusion Criteria

Since the aim of this study was to predict cardiovascular endpoints, patients who died of non-cardiovascular diseases (363 patients) were excluded, except, if they died after a period of more than 10 years (10 patients), as these patients can be considered as 10-year survivors for the purpose of this study. This left 2953 patients for inclusion in this study.

### Endpoint, subgroups and risk algorithm

The endpoint was defined as cardiovascular death due to cardiac causes (myocardial infarction, sudden cardiac death, death due to heart failure, death after intervention to treat coronary artery and other deaths due to coronary artery disease) and stroke. The study population was further stratified into patients with and without significant CAD. Thereby, significant CAD was defined as a lumen narrowing of 50 % or more in at least one of the coronary segments or a history of definite myocardial infarction.

CVD risk prediction in patients without CAD was compared with the CVD risk assessment tools recommended for asymptomatic persons, including the ESC Score (6), Framingham Risk Score(7), and the PROCAM Risk Score (8). In patients with CAD, the CV risk prediction was compared with the Marschner risk score (9) which was validated in patients with high to very high risk. In addition to qualitative comparisons of the existing risk scores, the category free net reclassification improvement, NRI(>0), and integrated discrimination improvement was used for comparison in patients without and with CAD (22)

### Principal Component Analysis of metabolic markers

For the Principal Component Analysis (PCA) the data (2953 patients) was normalized and adapted as follows: Outliers above the 99^th^ percentile and below the 1^st^ percentile were replaced by the respective percentile value. This mainly adjusts extreme outliers, which we assume to be due to technical issues. The adjusted data were translated to a 0-1 scale, with 0 representing the minimum and 1 the maximum value. This was necessary to allow comparisons of weights of the principal components to determine the influence of any specific variables. The subjects were then grouped into four age groups: 35-45, 45-55, 55-65, 65-100 years. Within each age group the median patient (i.e. medians for all variables) was calculated for both survivors and deceased patients. The PCA was performed using these median patients. To reduce the complexity any loading except the top five loadings (highest absolute loadings) were set to zero for plotting.

### Machine Learning Workflow

The overall processing workflow is visualized in figure 1. To generate and evaluate the model predictors, the input data was randomly split into a training and a validation dataset, the latter containing a random selection of 10% of the deceased patients, and an equally sized random set from the surviving patients. The deceased patients were selected such that the distribution of survival times was approximately the same as the distribution of survival times in the training data. The remaining data was weighted according to the class sizes, to address the imbalance, and the weighted data was used to train the models. To estimate the quality of the generated models, we decided to run our workflow 200 times, using different random selections for training and validation sets in each run. This allows us to provide a lower boundary for the quality of the generated models and gives an indication of the dependency of the prediction on the selected datasets. All results shown for the machine learning algorithms are the means of 200 runs (with added standard deviations, where applicable). The input data were all patients and the subgroups of patients without and with CAD as detailed above.

**Figure 1.**
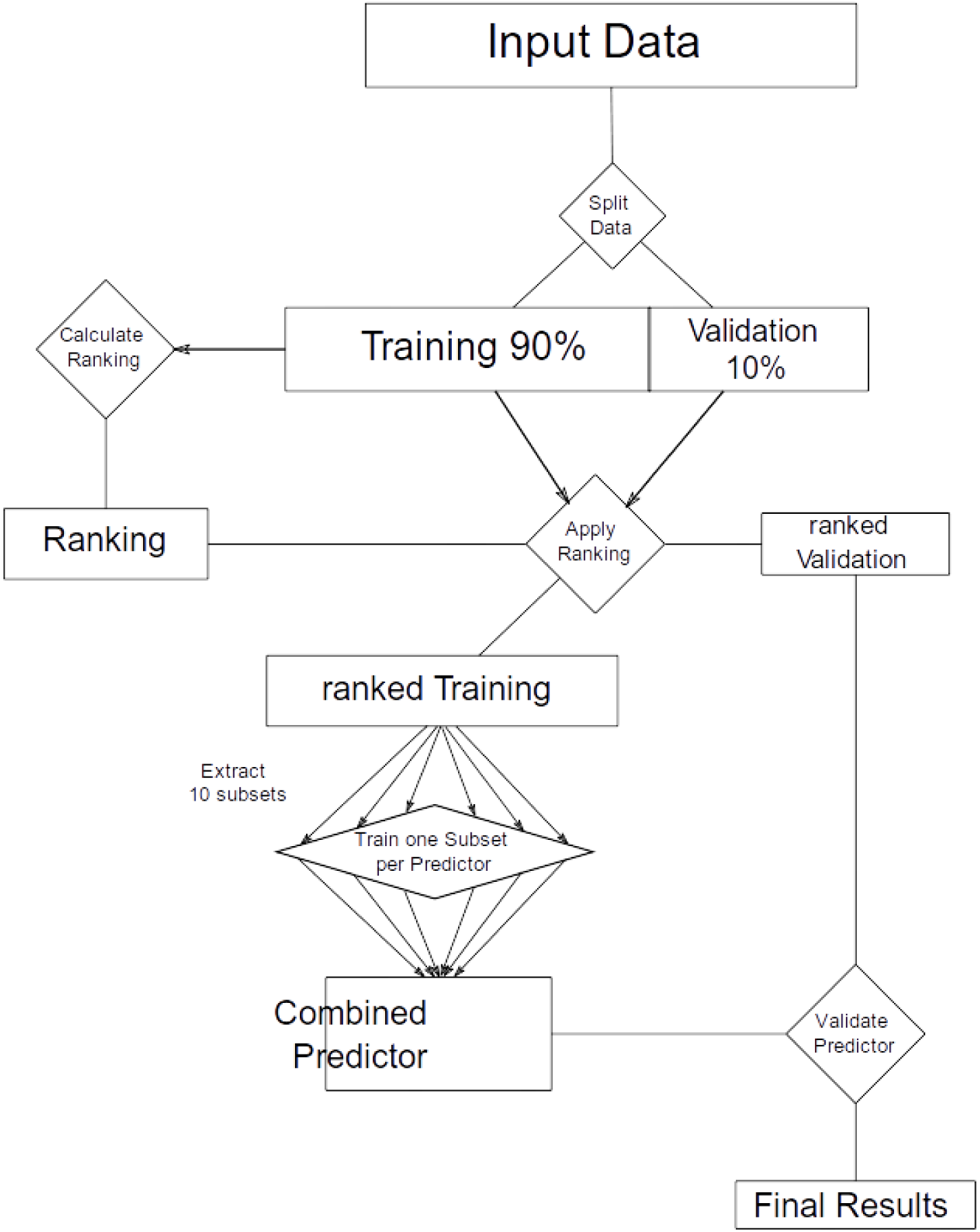
Overview of the predictor generation process. The upper part illustrates the attribute ranking which is performed on the whole data set. The data is then split into validation and training sets and the predictors are trained with data from the training set. For each predictor type 10 predictors are generated each using 30% of the training data from both survivor and dead classes. The final predictor is then built by averaging the results of the individual ten predictors and validated using the validation data.

### Ranking plasma and clinical markers

In general, a higher number of markers improves predictive power. However, it can similarly lead to overfitting if too many variables are taken into consideration, i.e., the patients/variables ratio becomes too small. We therefore tested the effect of the number of markers selected on the prediction accuracy and evaluated the improvement based on the Akaike Information Criterion (AIC) (16). To select the markers, we used three different ranking methods available in the WEKA library (17–19). The chosen methods were Correlation, information gain and information gain ratio (20). The ranking was performed in replicates of balanced sets to address the imbalance of classes, i.e., several survivors equal to the number of deceased patients were randomly selected from the survivors’ subgroup. 1000 rankings were conducted and combined using the sum of ranks of a marker from each ranking as score. The lower the final score the higher the marker was ranked in each ranking. To test the effect of increasing marker set sizes, as detailed above, we generated marker sets of up to 30 markers for each ranking method and used them as input to the model generation.

### Generation of prediction models

Multiple prediction methods were tested on each generated marker set. These included logistic regression, a polynomial and a radial basis function support vector machine, random forest and random tree methods as well as a naive Bayes predictor and a rule-based predictor as implemented in the WEKA library (17, 20, 21). For each ranking method, models were constructed based on the top ranked markers. Based on the respective marker set, ten sub-models were generated, each trained using 30% of the processed training data. The final class prediction was performed by averaging the predictions from the individual sub-models. The combination of multiple models allows us to avoid overfitting to individual samples, which is an issue given the relatively small number of samples in the study.

### Creation of RISK estimators

Since the machine learning algorithms used build models that determine a probability of a specific patient belonging to a particular class, our approach to create a risk estimator is based on this probability. In detail: We first obtained the probabilities for CV mortality from the respective predictor for each patient. Patients are then sorted by ascending probability. Each patient is weighted according to the outcome status. Due to the underrepresentation of patients with a cardiovascular (CV) death, any patient belonging to that category has a larger weight than a patient without a fatal event. The patients are then divided into ten equally weighted groups, in the order of their presence on the sorted list. This leads to groups of less patients, when there is a higher percentage of CV deaths, and to groups with more patients, if there is a lower percentage of CV deaths. For each of the groups, we then calculated the percentage of members in the group who died from CV events and calculated the average of the group’s minimum and maximum probability as determined by the model. These average and percentage combinations were used as anchor points to fit a quadratic curve that was then used as a risk estimator using Apaches Commons™ - Math curve fitting tools. Other types of curves (exponential and linear as well as higher order polynomials), where also tested, but a quadratic model yielded the best fit. A more detailed explanation can be found in the Supplemental Data (Table S1, S2 and Figure S1).

### Implementation

The prediction framework was implemented in Java building on the WEKA library (17). The evaluation scripts were implemented in Matlab. Statistical analyses were performed in SPSS 25.0 statistical package (SPSS Inc., Chicago, IL, USA) and R 3.4.1. The code is available upon request.

## Results

### Initial principal component analysis

First, we conducted a simple PCA analysis using the blood plasma biomarkers (see figure 2). We observed a good separation of the median values of survivors and deceased patients from different age groups. This separation was the stronger the younger the patients were, and grew weaker in older patients. We also found that individual patients aged below 50 separated well on this PCA.

**Figure 2.**
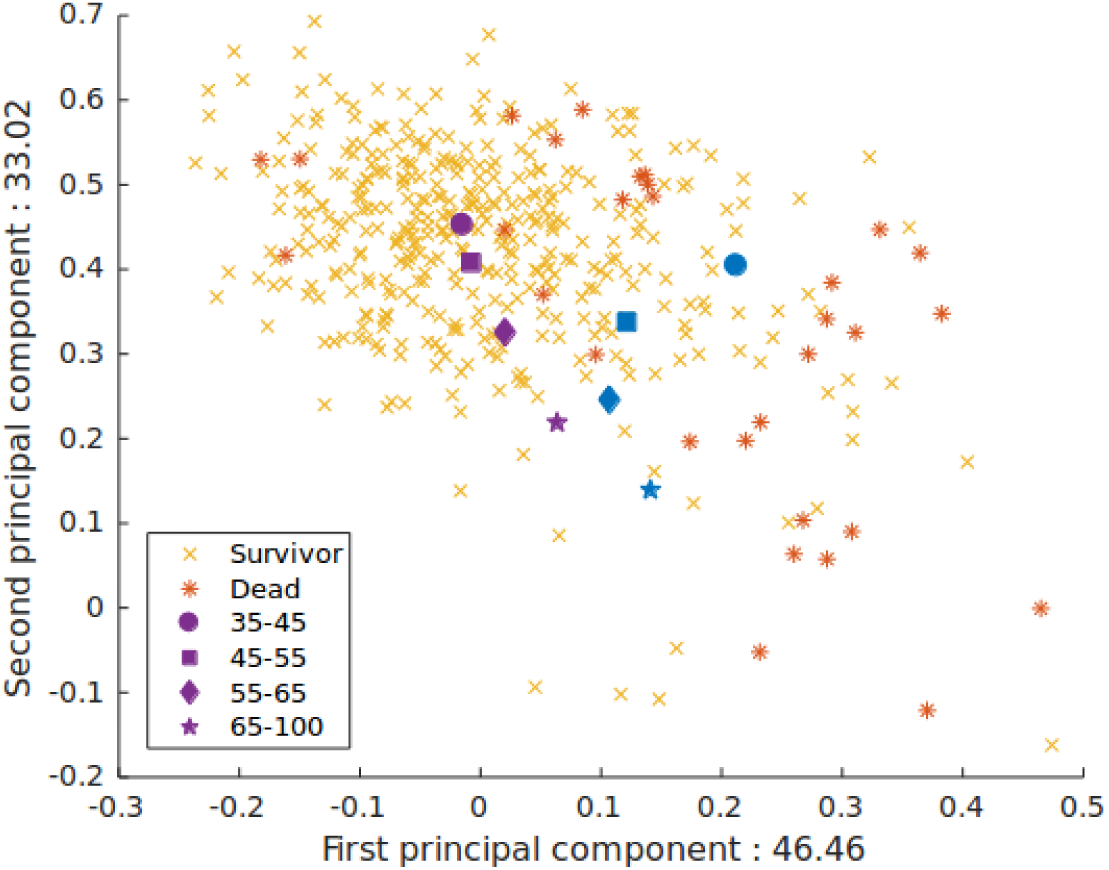
Principal component analysis using the medians of four different age groups (35-45, 45-55, 55-65, and 65-100). Purple: Survivors medians, Blue: Deceased personś medians. The medians separate well. The individual patients plotted are patients aged below 50 (orange x – survivors, red stars - deceased), for whom a good separation is achieved. Interestingly, the differences in the medians between survivors and deceased drop with higher age.

The results of the PCA indicated that we could indeed find separating properties even with a small number of variables in an unsupervised hypothesis-free way. On the basis of these initial investigations, we decided to test multiple supervised machine learning techniques to generate models for survival and to establish risk estimators.

### Risk of CV mortality overall

Baseline characteristics of the overall study population are given in supplementary table S3. 603 patients deceased of CV causes, whereas 2350 patients survived within ten years of follow-up. Patients who deceased were predominantly male, significantly older and had higher systolic blood pressure, more diagnosed diabetes mellitus type II and a history of previous myocardial infarction. Patients who died had significantly more severe CAD (more than 50 percent lumen narrowing in three vessels) whereas more patients who survived had more often no or less CAD.

We tested three different ranking schemes in combination with seven different predictors and calculated for each approach the accuracy, AIC and area under the curve (AUC). Overall, we found that the InfoGain ranking scheme provided the best results (supplemental figure S1 A and B). Using this ranking scheme we obtained AUC values for the selected prediction algorithms with the best result obtained for the Random Forest predictor with an average AUC of 0.78 (±0.05), followed by the random tree (0.77±0.05), the rule based approach (0.77±0.05), logistic regression (0.76±0.05) and the radial basis function SVM with 0.76±0.05. Overall, the different machine learning approaches generated qualitatively very similar predictors and we therefore decided to use a selection of these for further comparison. Since random trees are a subclass of random forests we decided not to use it for further analysis and restricted our further work to the remaining four (diverse) methods.

The ranking results of the first 30 markers according to the InfoGain scheme is shown in supplementary figure S2 A. We found that using more than four predictors leads to a comparatively high increase in effort and cost (based on the Akaike Information Criterion - AIC), without further improving the accuracy of the generated models^14^. This can also be observed in supplemental figure 2 B, which shows an increasing AIC for larger marker counts whereas no gain in accuracy was obtained (also shown in supplemental figure 1).

Based on this selection criterium the four highest ranked markers with the strongest association to cardiovascular mortality were: NT-proBNP (N-terminal pro-BNP), followed by TNT (Troponin T), GFR (estimated glomerular filtration rate) and age.

To determine the discrimination power we performed average Kaplan-Meier plots of the four predictors (logistic regression, random forest, rigor rule based, support vector machine) which produced good separations between survivors and deceased (supplementary figure S3).

The results of our workflow with a combination of InfoGain ranking and four different predictors are given in supplementary figure S4. All predictors have assigned more deaths to the high-risk group and more patients who have survived were assigned to the low-risk group. In general, there were no significant difference between each predictor.

### Risk of CV mortality in patients without CAD

Baseline characteristics of the subpopulation without CAD are given in supplementary table S4. In the subpopulation without CAD 95 patients deceased and 761 survived for at least ten years. Patients who died were again significantly older, had a lower left ventricular ejection fraction (LVEF), had more diagnosed diabetes mellitus type II and had higher degrees of lumen narrowing 11-59%. No patient had a previous myocardial infarction or severe coronary vessel disease (defined as lumen narrowing > 50% in one or more coronary vessels).

We observed that the InfoGain ranking still yielded the best results (supplementary figure S5). Again, using four markers providing good AUCs (0.835±0.1014 for random forests). The markers which were ranked highest were equal to the obtained ranking in the overall population. From highest to lowest: NT-proBNP (N-terminal pro-BNP), followed by TNT (Troponin T), GFR (estimated glomerular filtration rate) and age (figure 3).

**Figure 3.**
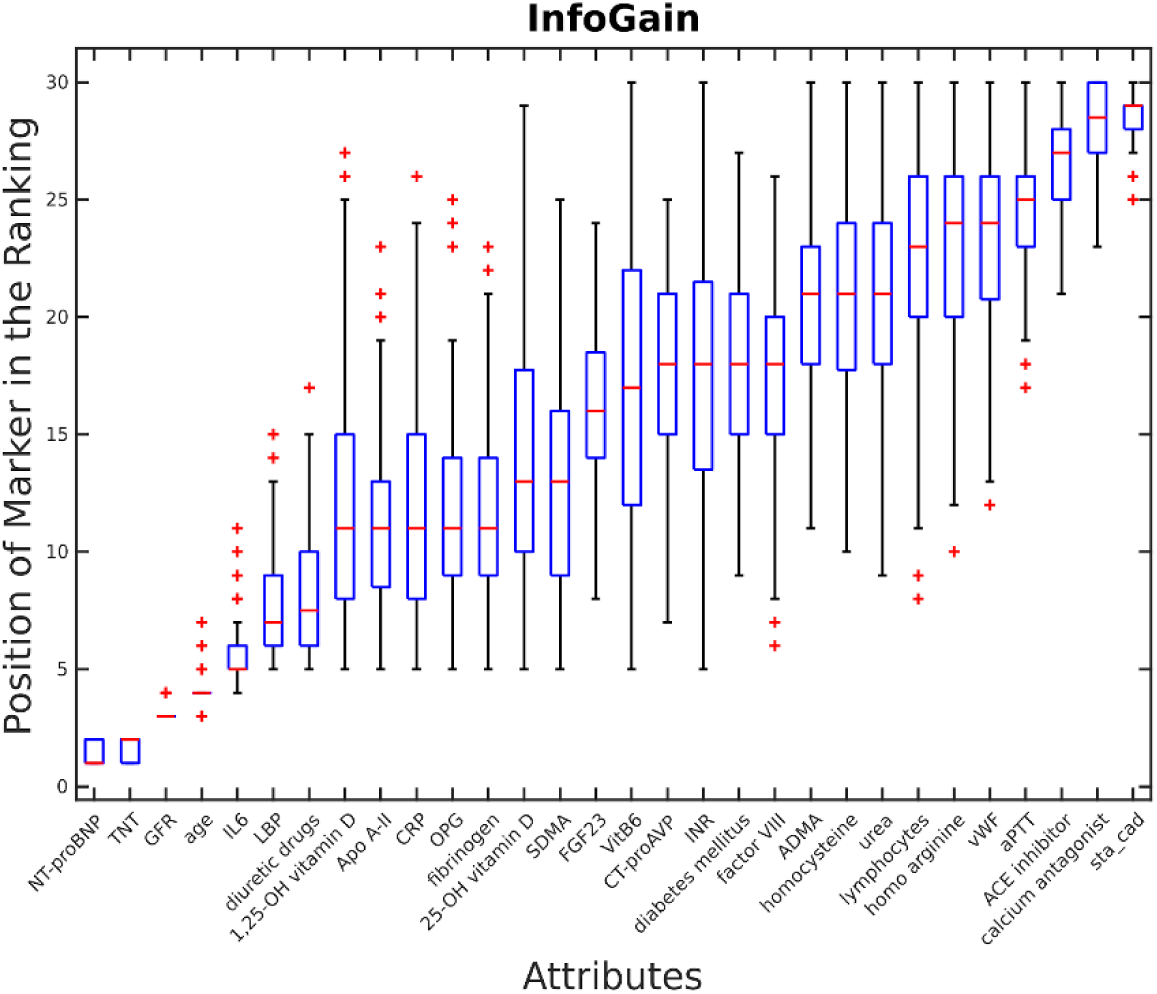
Ranking results of the first 30 markers by the info gain scheme in patients without CAD.

After creating a risk predictor based on the results of a combination of InfoGain ranking and four different predictors we compared the predicted risks with established risk assessment tools. (Figure 4). Overall, we observed, that the machine learning approaches assign a larger number of true at risk patients to the high risk group than existing scores (figure 4B). Simultaneously, the existing algorithms in general tend to assign more patients into the medium risk groups, while the machine learning algorithms tend to have a clearer risk/no risk assignment (figure 4B/D), reflecting a higher specificity of the machine learning approaches. This is to be expected, given that the machine learning algorithms were trained on a two-class prediction. Overall, the machine learning models trained in this study miss fewer at risk patients and are better in determining high risk patients compared to existing scores. This is also reflected in the NRI and IDI scores, when the model predictions are compared to ESC, FRAMINGHAM and PROCAM respectively. Here, the machine learning approach achieves a NRI(<0) of 0.86/0.91/0.85 and an IDI of 20.84/21.92/18.17 for ESC/FRAMINGHAM/PROCAM respectively.

**Figure 4:**
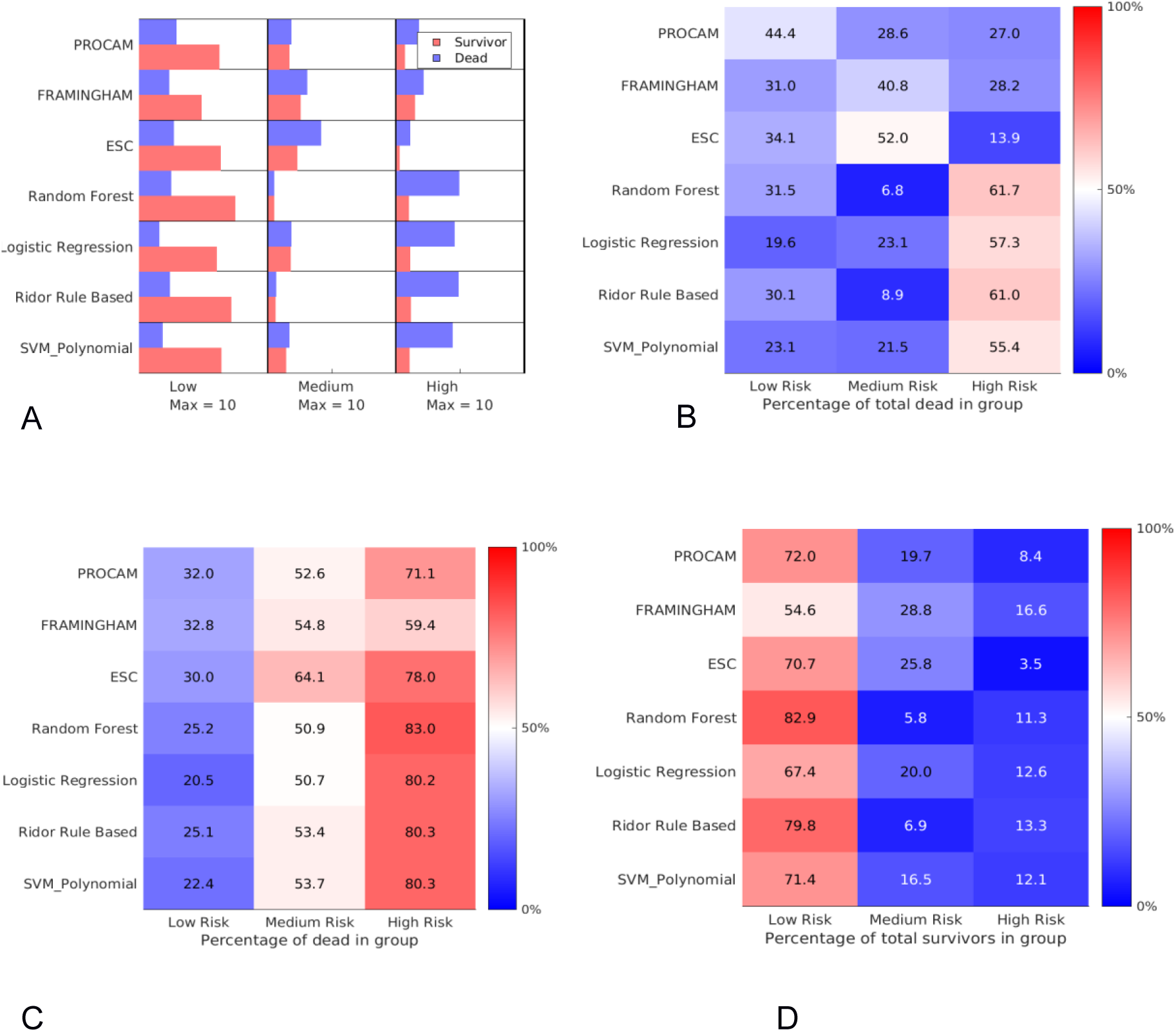
Comparison of established and generated risk predictors.CV mortality of patients without CAD. The risk estimators indicated low (<10%), medium (10-20%) and high (>20%) risk. All values are based on averages of the patients from the individual validation sets (8 dead and 9 surviving patients for each replication). **A** shows the average classification of patients by the scorers on the validation sets. For comparability the bars were adjusted such that all groups (low, medium, high) are scaled to the largest group in any of the predictors (here, a total of ∼10 patients assigned to the survivor group by the random forest predictor). **B** shows the average percentage of validation patients without CAD who died being classified into either low/medium or high risk by the predictors. **C** shows the average percentage of deaths of the validation patients who died classified to low/medium/high risk category by the predictors. **D** shows the average relative classification of validation patients without CAD that survived.

### Risk of CV mortality in patients with CAD

We wished to compare our method to the Marschner score for prediction in patients with higher degrees of CAD. Since the Marschner score predicts six years risk, we built a five years score from our data.

Baseline characteristics of patients with CAD are given in supplementary table S5. 247 patients with CAD deceased due to cardiovascular causes and 1618 survived within five years. Patients who died were significantly older, had more diagnosed diabetes mellitus type II and had a lower LVEF. Patients who deceased had more vessel disease compared to the survivors (3 VD vs. 1-2 VD).

The ranking of the markers is shown in supplementary figure S6. The four highest ranked markers were: NT-proBNP (N-terminal pro-BNP), GFR (estimated glomerular filtration rate), CT-proAVP (C-terminal pro arginine vasopressin) and TNT (Troponin T).

The resulting AUC at 0.79±0.05, using InfoGain and a random forest approach, is lower in comparison to the AUCs observed in patients without CAD (supplementary figure S7). Four markers provided the best tradeoff between cost and accuracy and we used those models for the comparison with the Marschner score.

Both the established Marschner score and our automatically generated score achieved qualitatively similar results, with a slight advantage of an average of 80% (present workflow) vs 75% (Marschner) of high-risk patients dying within five years (supplementary figure S8 C). Again, the automated scores showed a smaller number of patients assigned to a medium risk, while the Marschner score assigned 10-20% risk to almost 50% of the total population. The automatically generated risk predictors generate a more bimodal distribution of predicted risks (supplementary figure S8 A/B), with a higher percentage of survivors being classified as low risk compared to the Marschner score (supplementary figure S8 5D). However, the improvement, with respect to NRI and IDI is less pronounced, with an NRI of 0.57 and an IDI of 10.25.

Unfortunately, the general risk prediction method is not directly comparable to any existing risk score. Our outcome comprises the hard endpoint CVD death, while most risk scores predict a larger range of CVD-related endpoints, and scores, which predict probabilities of CVD death, are commonly restricted to subpopulations. For example, the FRAMINGHAM, ESC and PROCAM risk scores were designed to be applied to asymptomatic patients, while the Marschner score was built for patients with CAD. In contrast, our general score predicted CV death irrespective of the CAD status of the patient.

## Discussion

This study of long-term prediction of fatal CV events has several important findings. First, four or less laboratory markers are sufficient to predict long-term CV mortality in stable coronary artery disease patients. Second, machine learning techniques are superior to standard linear statistical models in prediction of long-term CV mortality. Finally, we present here several prediction models, for both five- and ten years prediction in patients without and with CAD, respectively, which shows that our approach can easily be applied to other research questions, and we provide the scripts used for model generation and evaluation online.

### Risk factors

We have selected our risk markers out of an array of 184 biomarkers and 21 clinical markers and ranked each marker on their contribution of causing CV mortality. The ranking was obtained by a combination of three different ranking schemes in combination with seven different predictors. For each approach the AUC, accuracy and AIC was determined and compared for the best prediction power. In the overall study population and the subpopulation without CAD we have found the same key set of clinical and laboratory markers as major indicators of ten-year CV mortality, which were ranked from the highest to the lowest as followed: NT-proBNP (N-terminal pro-BNP), followed by TNT (Troponin T), GFR (estimated glomerular filtration rate) and age. The markers differed slightly for patients with CAD where NT-proBNP, GFR, CT-proAVP and TNT were the highest predictive markers for 5-years CVD mortality. We found that markers associated with hemodynamic status such as NT-proBNP and CT-proAVP were higher ranked in patients with CAD. In general, renal dysfunction was consistently associated with CVD mortality, in the overall and each subpopulation. Surprisingly, with the exception of age, there were no clinical or anthropometric variables among the top-ranked predictors in each of our models. Our data are in accordance with a previously published study which examined 30 novel biomarkers in a population cohort with 538 incident cardiovascular events and 10-years follow-up (23). The strongest associations were found for NT-proBNP, C-reactive protein, and sensitive troponin I from which a biomarker score was developed.

In consequence, we found that risk factors differed across the subpopulations explored and time periods. Therefore, we suggest that the ranking of the markers is a critical step before training a predictor. In addition, a fixed reduced set of markers makes overfitting less likely. We postulate that the InfoGain ranking scheme achieved the best results for all tested predictors and we further suggest that age, the estimation of GFR, a main cardiac markers like TNT and a hemodynamic marker such as NT-proBNP are sufficient in prediction of overall CV-mortality.

### Comparison to existing Risk Scores

We have compared our risk prediction model with existing risk assessment models, including the FRS, PROCAM and ESC-Score.

First, all machine learning algorithm have allocated patients who have died to a higher risk group in comparison to the conventional risk scores. Vice versa, patients who have a high risk are more likely being classified in a high risk group by a machine learning algorithm than by a conventional risk score and might be referred more likely to a further treatment.

Second, patients who have survived have been allocated more often into a lower risk category by machine learning algorithm in comparison to the other examined risk scores. Hence, a patient who has a low risk is more likely classified in a low-risk group than by another risk score. We therefore postulate that risk assessment based on machine learning methods is preferable over risk models derived from conventional statistical methods.

In general, the machine learning algorithm achieved a higher reliability by classifying survivors more frequently as low and deceased patients more frequently as higher risk persons. The percentage of patients assigned to the medium risk category was lowest for the machine learning algorithms.

It is important to note that e.g. PROCAM predicts a different outcome (both myocardial infarction and death due to CVD) than the outcome used in this study (death due to CVD). The same is true for the FRS, which predicts the risk of myocardial infarction. Therefore, the classification according to risk is skewed in these two predictors, leading to a higher number of patients being classified into the medium category. However, considering that the classifications of FRS and PROCAM would shift many patients to the survival groups, it is obvious that this would lead to a very small number of patients being classified to the high risk group, indicating that the models suggested in this study are more likely to detect patients who are at risk.

The only risk score directly comparable to our models is the ESC score, which had the same end point, and which clearly shows a weaker ability to distinguish between high and medium risk patients.

However, comparing machine learning algorithm with the Marschner score in patients with CAD has shown equivalent results. The Marschner Score has classified patients as good as the developed machine learning algorithm.

In conclusion we conclude that the combination of an InfoGain ranking scheme with a random forest predictor has performed the best. Our proposed prediction scheme has assigned patients accurately in their true risk group, achieved the highest sensitivity and accuracy independently if deaths or survival is predicted and further independently of the subpopulation and time period that is observed.

### Benefits of dynamic risk assessment

In the past decades CVD risk assessment has been realized by the development of risk charts and calculators. Each risk score estimates CV risk using a different set of markers, different time frames and clinical endpoints. A comparison of 25 different risk calculators in 128 hypothetical patients showed that the risk categories agreement between pairs of calculators was only 67%. Further, the pairs of calculators which assigned a different category to the same patient were approximately one third (12). Furthermore, few of the risk calculators were insufficiently validated which may yield problems when the risk assessment model is applied to a population substantially different from the study cohort (10).

The comparison of CV death rates across European countries reveals a substantial variation (24, 25). The highest CV mortality burden is found in central and eastern European countries compared to Northern, Southern and Western countries. While genetic and environmental factors, such as nutrition and lifestyle, have a high impact on CV disease we have developed a dynamic model, which can directly be adapted to each study. We have achieved this goal by creating a CV risk prediction model which is scoring according to the contribution of CV mortality in the population. Further the model is comparing different risk prediction algorithms with each-other in multiple runs and offers the best prediction algorithm for a distinct population. Overfitting is avoided by multiple runs in a ten-fold cross validation approach. Further, our approach offers the possibility to estimate the overall quality of the prediction based on the robustness of the performed runs. We anticipate, that when training the predictors on the whole data set our results would improve or at least stay within the quality presented here.

### Limitations

Many analysis techniques do not cope well with missing data. Other methods can commonly infer the missing data by imputation. To achieve a consistent way how missing values are treated, we replaced missing values in all datasets by the median values of the available values.

Samples have been drawn in patients initially presenting to a tertiary cardiac center for coronary angiography and some laboratory values might be elevated by increased emotional stress prior to examination or ischemic heart disease. Subsequent treatments, procedures and discharge medication may have influenced our long-term mortality rates. Further, we have focused on fatal-CV events only.

The patient cohort used might have influenced the effectiveness of other risk predictors, and thus present a bias towards the generated scores. However, since the main aim of this study was to establish an easy protocol for risk predictor generation, we expect that our results would hold in other studies if the found markers are determined in those studies.

## Conclusion

We have developed a CV prediction model based on machine learning techniques using a comprehensive database of clinical, routinely and non-routinely measured laboratory data. The machine learning algorithm achieved a higher reliability by classifying survivors more frequently as low and deceased patients more frequently as higher risk persons. The percentage of patients assigned to the medium risk category was lowest for the machine learning algorithms in comparison to the other examined risk scores. Further, we created a fully automatic and self-validating framework, which is easily applied to a broad spectrum of populations, clinical endpoints and time periods of follow-up and made this framework available online.

## Data Availability

All data produced in the present study are available upon reasonable request to the authors.

## Acknowledgments

We extend our appreciation to the participants of the LURIC study. We thank the LURIC study team which was either temporarily or permanently involved in patient recruitment, sample, and data handling. We also would like to thank the laboratory staff at the Ludwigshafen General Hospital, Universities of Freiburg, Heidelberg, Ulm (Germany) and Graz (Austria).

## Supplementary Data

### Description of the Risk estimator generation

Assume that we have 1000 patients. 300 off those are in the CVD dead group and 700 survived. The trained predictors assign a probability to be in the CV death group to each patient. In addition. a weight is assigned to each patient according to whether they belong to the majority (survivor) or minority group. The majority group gets a weight of one. while the minority group gets a weight of #MajorityGroup/#MinorityGroup.

---

**Supplement Figure S1.**
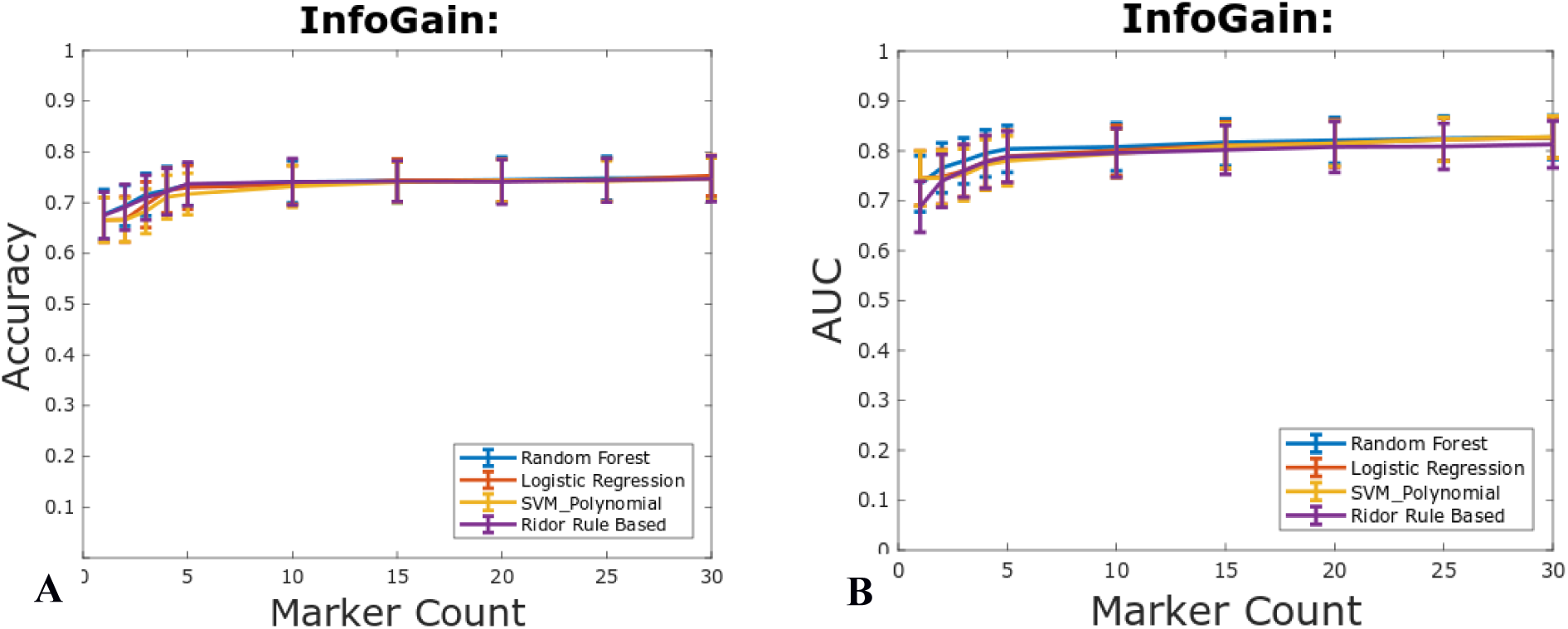
Accuracy and area under the curve (AUC) of the three tested ranking schemes in combination with seven different predictors in the overall study population. The InfoGain ranking scheme and four predictors yield the best results.

**Supplement Figure S2.**
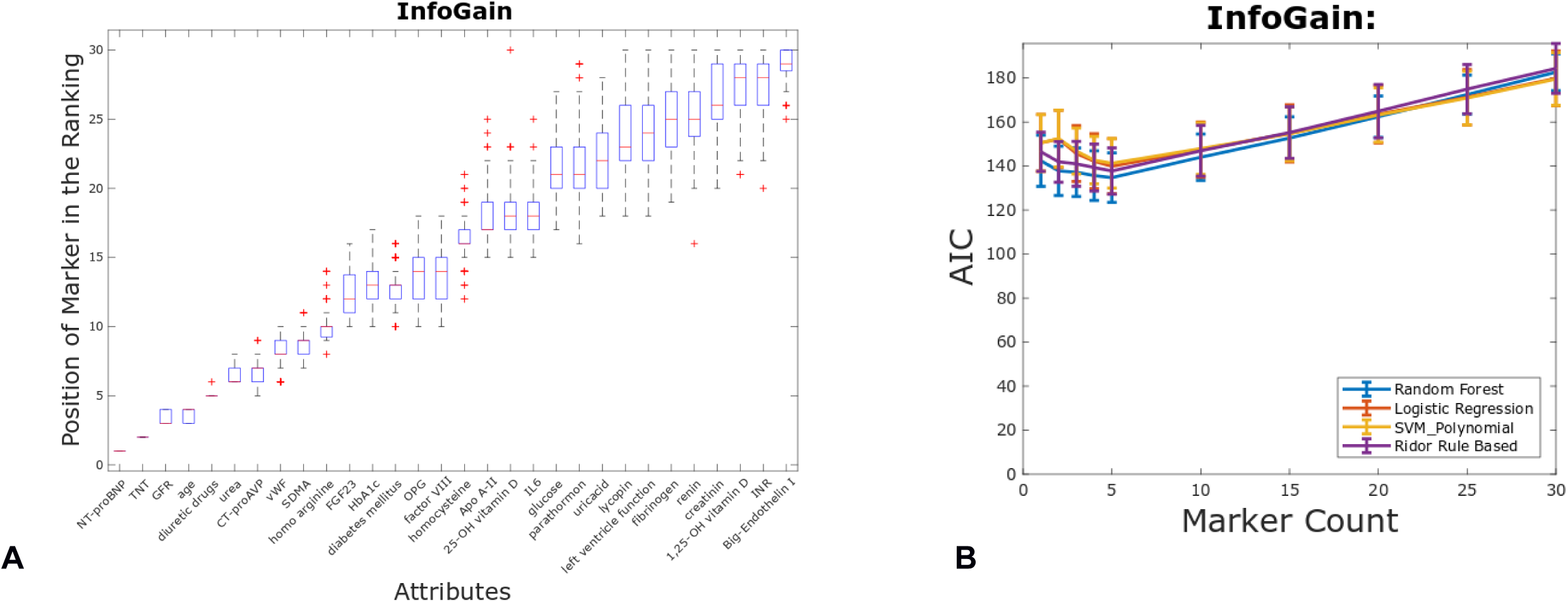
**A.** Ranking results of the overall study population of the first 30 markers by the info gain scheme. **B.** Akaike Information Criterion (AIC) increased with increased marker taken into the model.

**Supplement Figure S3.**
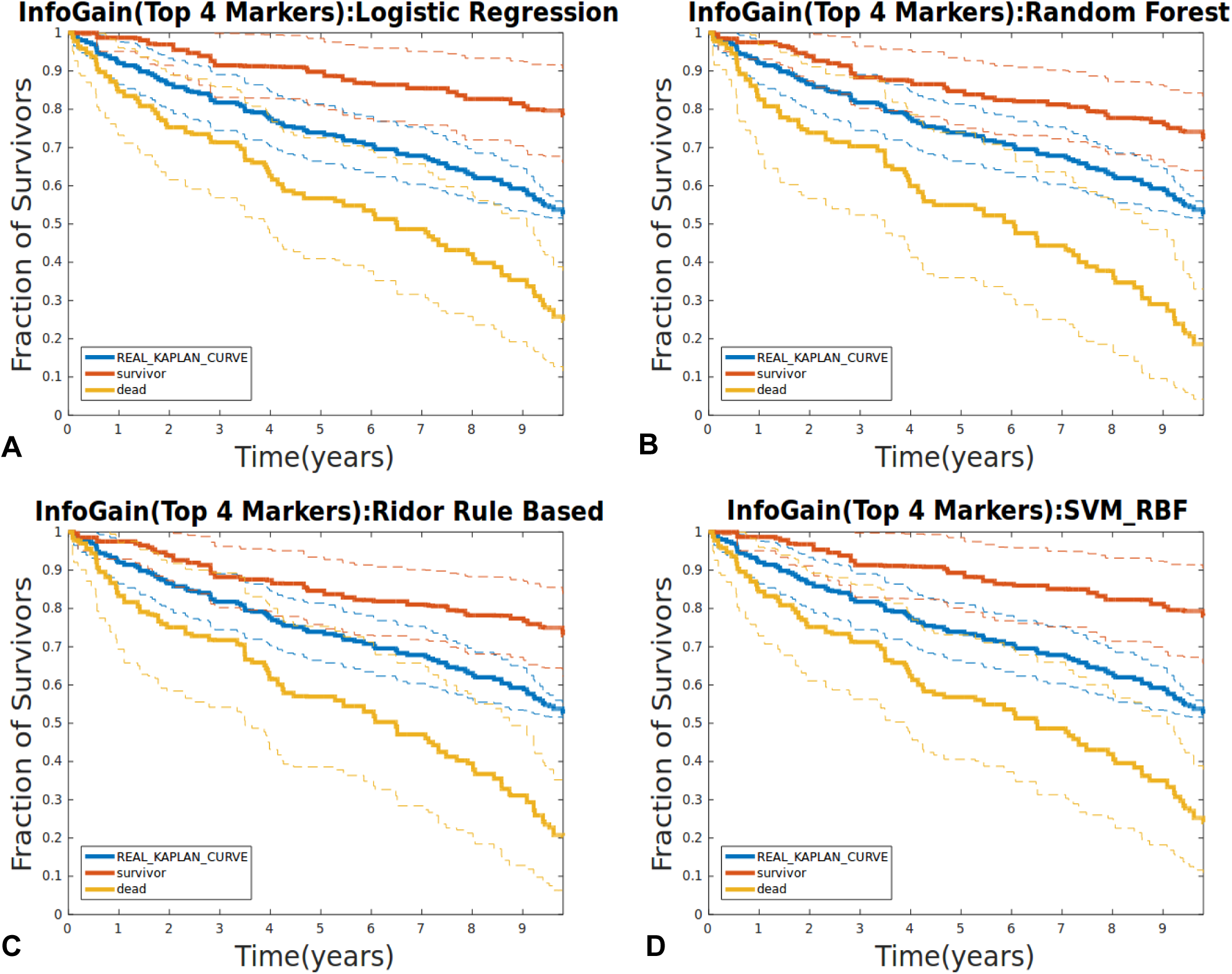
Kaplan-Meier plot of the patients predicted for CV mortality and survivors in the overall study population. The predictors used are **A.** logistic regression; **B.** Random Forest; **C.** Rigor rules and **D.** Support vector machine (radial basis function kernel). The dashed lines indicate the standard deviations at each potential event time. The ‘REAL_KAPLAN_CURVE’ represents the population curve not divided into classes and is shown for comparison.

**Supplement Figure S4:**
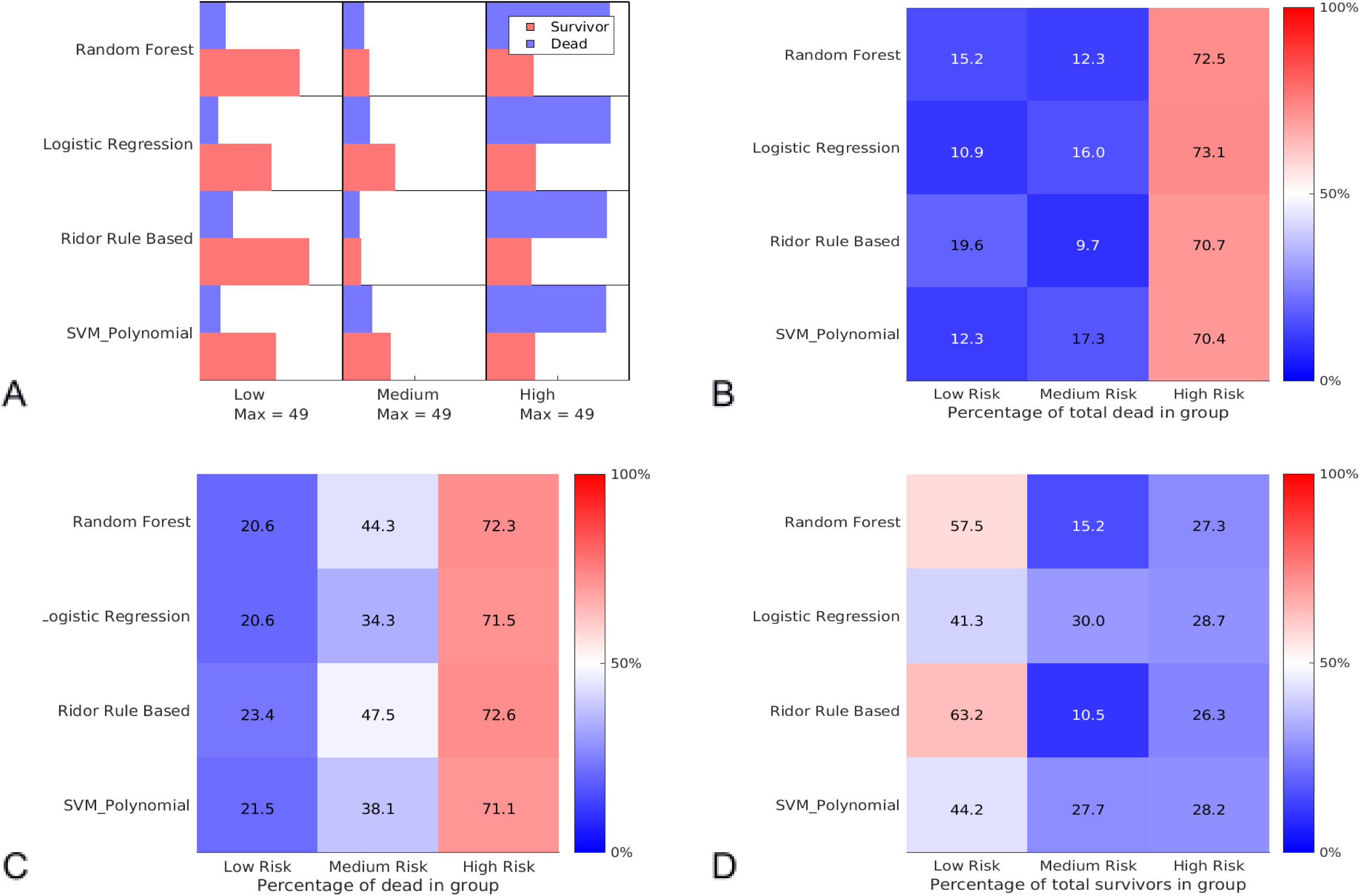
Comparison figure for the general risk scores generated on the complete data set. The risk estimators indicated low (<10%). medium (10-20%) and high (>20%) risk. All values are based on averages of the patients from the individual validation sets (21 dead and 21 surviving patients for each replication). **A** shows the average classification of patients by the scorers on the validation sets. For comparability the bars were adjusted such that all groups (low. medium. high) are scaled to the largest group in any of the predictors (here. a total of ∼23 patients assigned to the survivor group by the random forest predictor). **B** shows the average percentage of validation patients with CAD who died being classified into either low/medium or high risk by the predictors. **C** shows the average percentage of deaths of the validation patients who died classified to low/medium/high risk category by the predictors. **D** shows the average relative classification of validation patients with CAD that survived.

**Supplement Figure S5.**
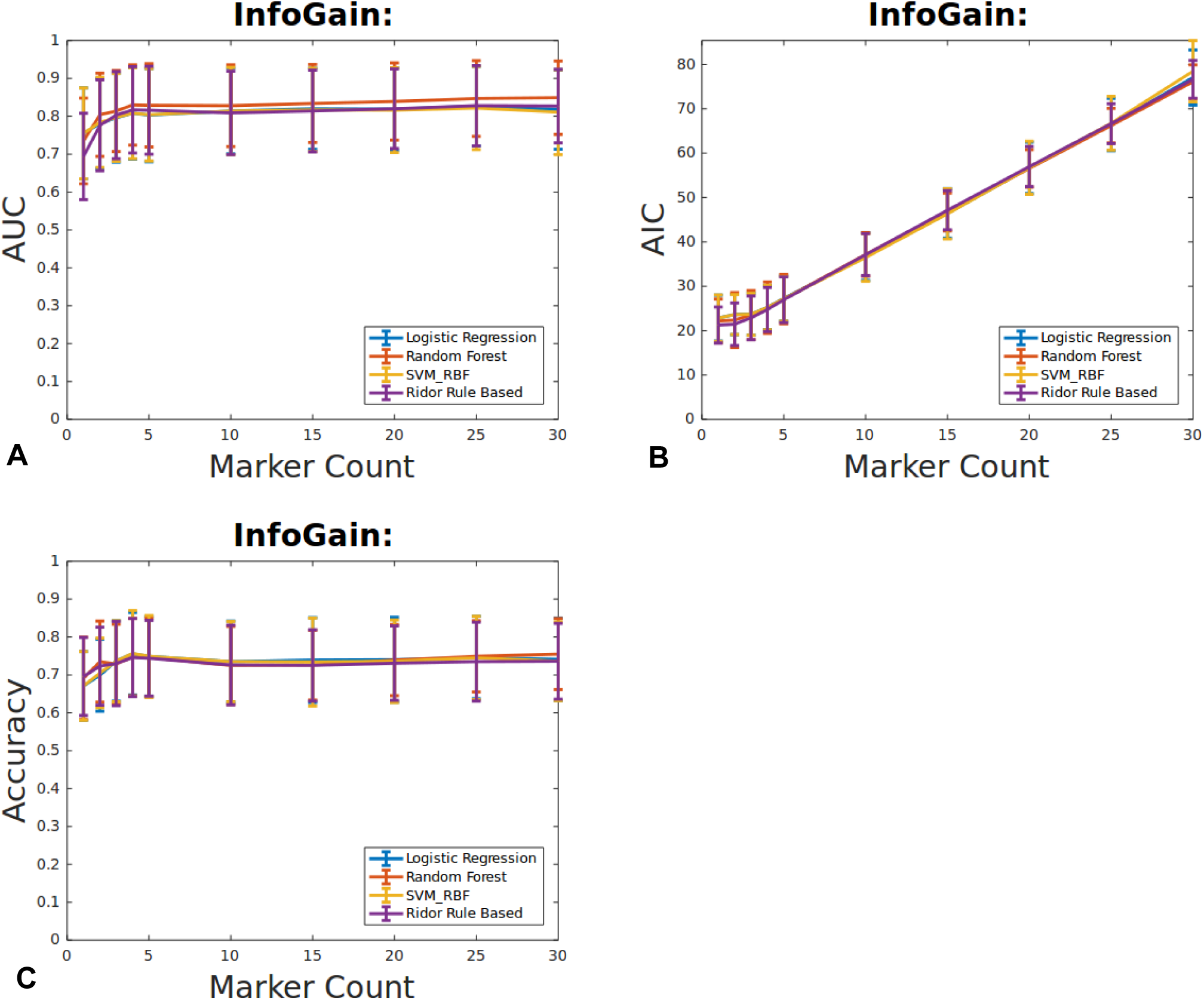
Area under the curve (AUC). Accuracy and Akaike Information Criterion (AIC) of the three tested ranking schemes in combination with seven different predictors in patients without CAD. Ranking according to the InfoGain scheme.

**Supplement Figure S6.**
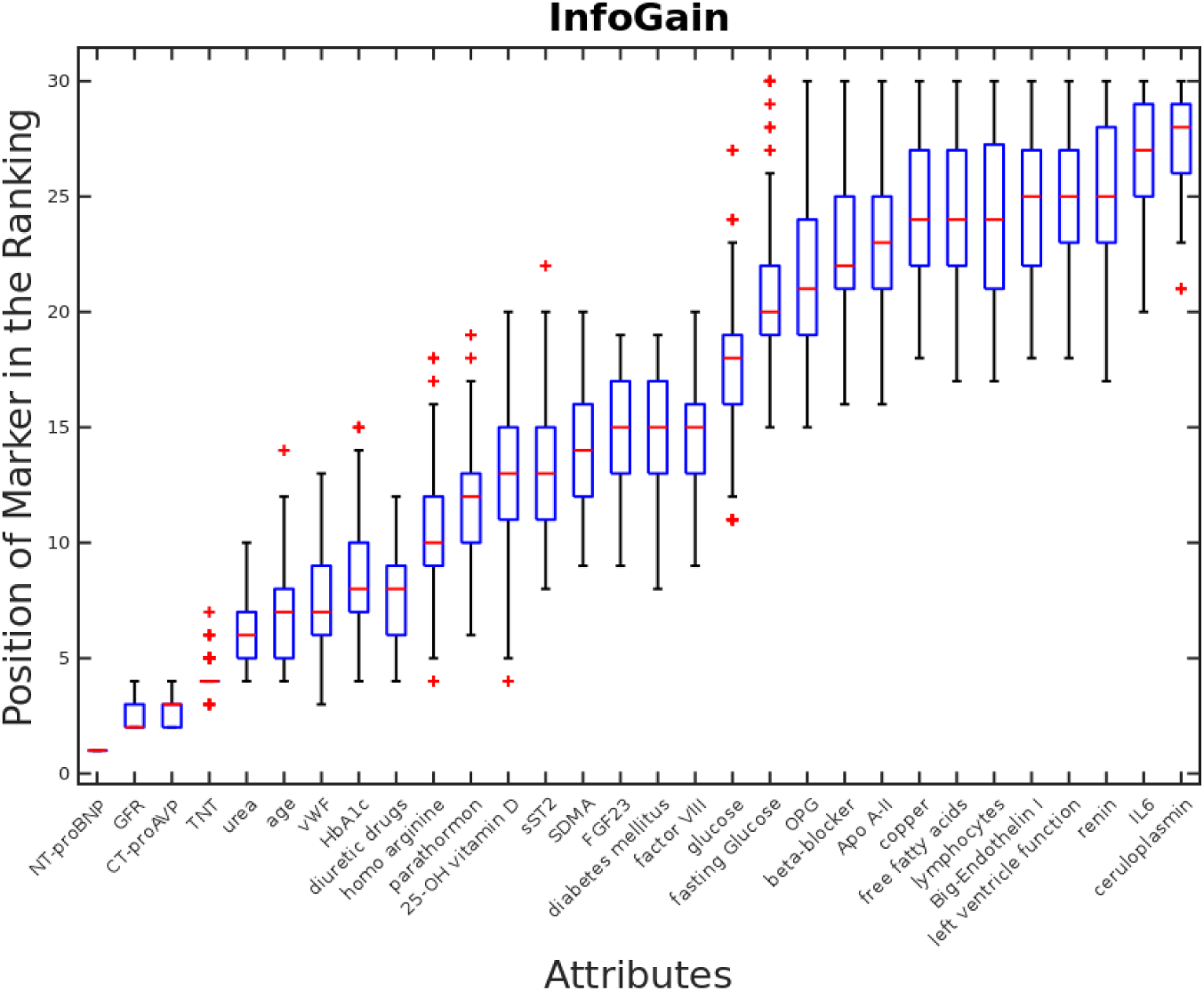
Ranking results of the first 30 markers by the info gain scheme in patients with CAD.

**Supplement Figure S7.**
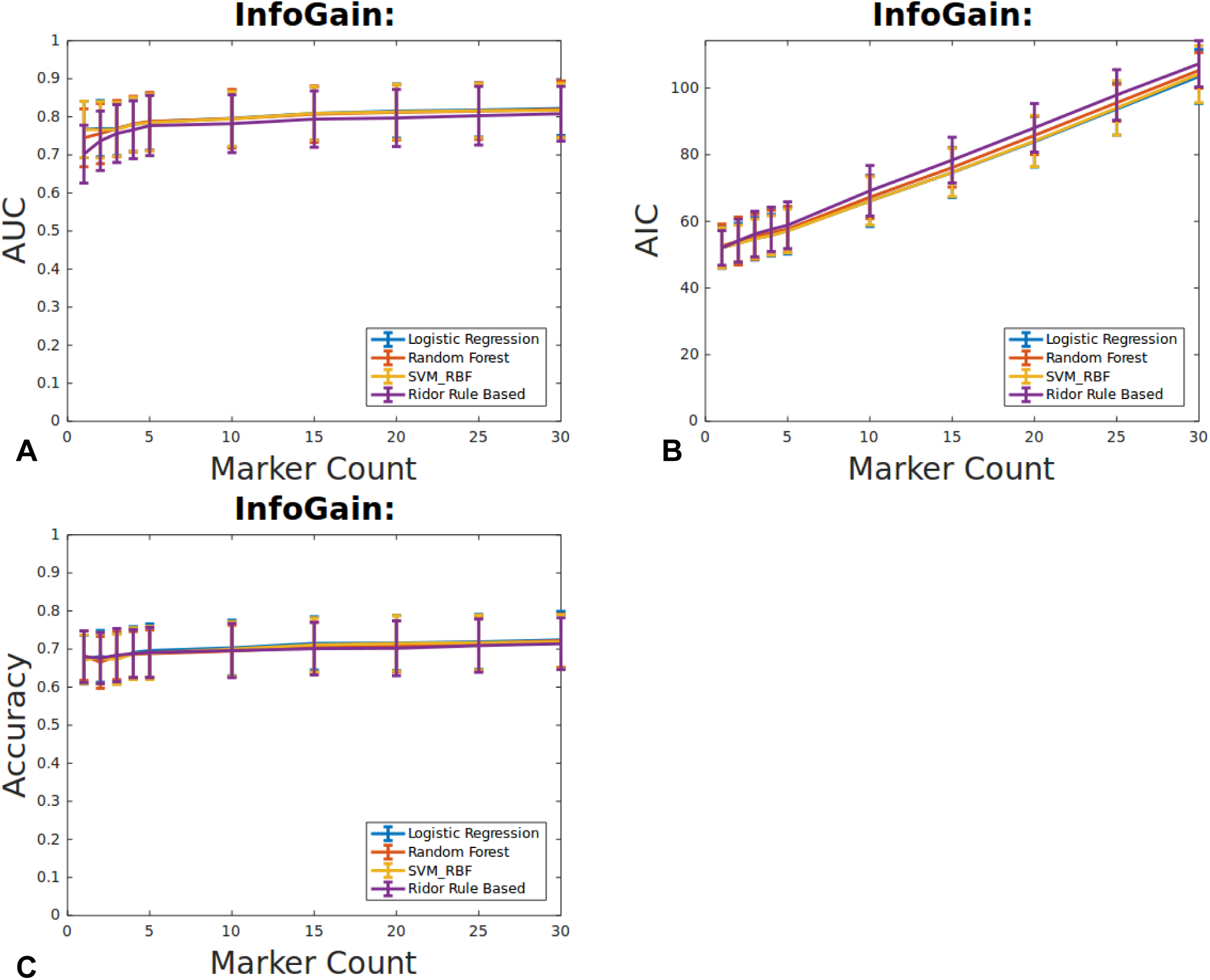
Area under the curve (AUC). Accuracy and Akaike Information Criterion (AIC) of the three tested ranking schemes in combination with seven different predictors in patients without CAD. Ranking according to the InfoGain scheme.

**Supplement Figure S8:**
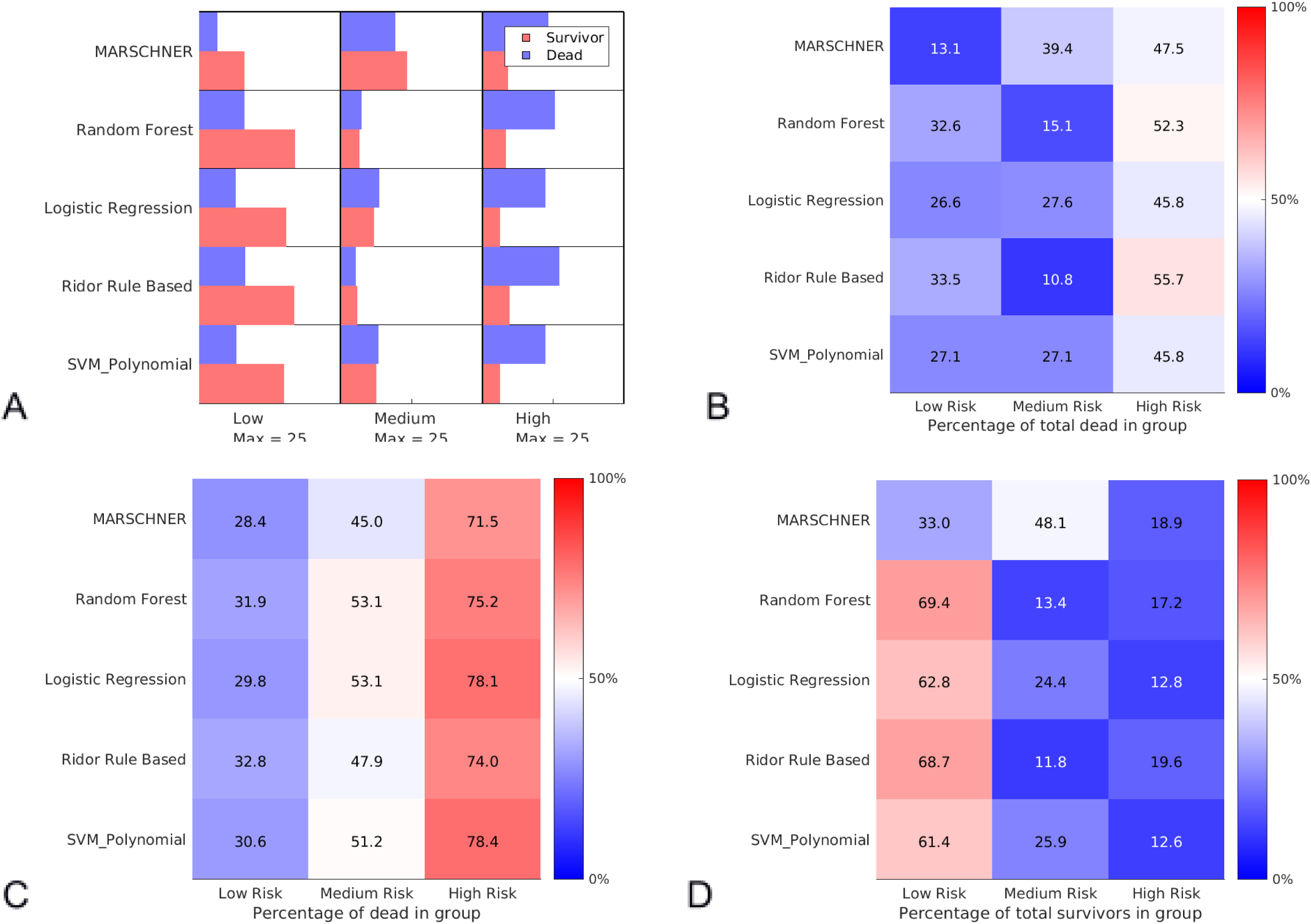
Comparison figure for the risk scores generated in patients with CAD. The risk estimators indicated low (<10%). medium (10-20%) and high (>20%) risk. All values are based on averages of the patients from the individual validation sets (21 dead and 21 surviving patients for each replication). **A** shows the average classification of patients by the scorers on the validation sets. For comparability the bars were adjusted such that all groups (low. medium. high) are scaled to the largest group in any of the predictors (here. a total of ∼23 patients assigned to the survivor group by the random forest predictor). **B** shows the average percentage of validation patients with CAD who died being classified into either low/medium or high risk by the predictors. **C** shows the average percentage of deaths of the validation patients who died classified to low/medium/high risk category by the predictors. **D** shows the average relative classification of validation patients with CAD that survived.

**Supplement Table S1.** An example of assigned probabilities and weights.

|  | P2 | P3 | P5 | P6 | P7 | P8 | P9 | P10 | P11 | P12 | P13 | P15 | P16 | P17 | ... | PN |
| --- | --- | --- | --- | --- | --- | --- | --- | --- | --- | --- | --- | --- | --- | --- | --- | --- |
| P(CVD) | 0.25 | 0.77 | 0.41 | 0.86 | 0.14 | 0.21 | 0.12 | 0.01 | 0.87 | 0.36 | 0.92 | 0.15 | 0.36 | 0.41 |  | 0.86 |
| Class | CVD | S | S | CVD | S | S | S | S | CVD | S | CVD | S | CVD | S |  | S |
| Weight | 7/3 | 1 | 1 | 7/3 | 1 | 1 | 1 | 1 | 7/3 | 1 | 7/3 | 1 | 7/3 | 1 |  | 1 |

**Supplement Table S2.**
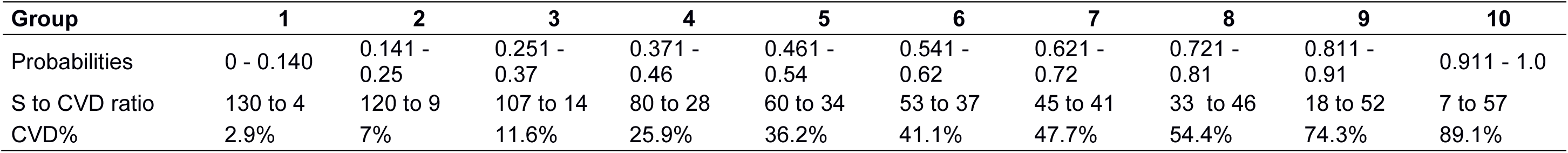
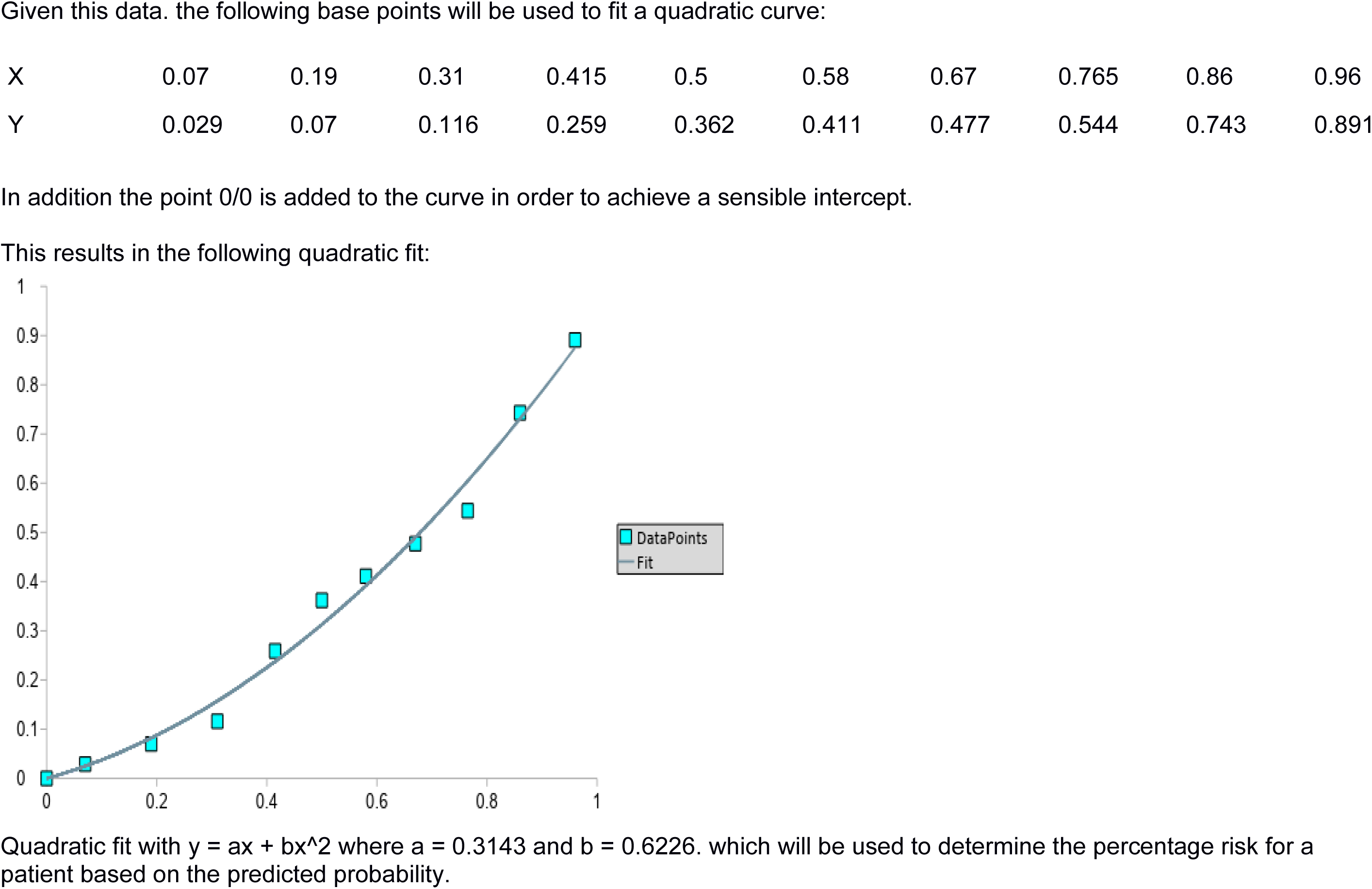
Assumption of the distribution

| Group | 1 | 2 | 3 | 4 | 5 | 6 | 7 | 8 | 9 | 10 |
| --- | --- | --- | --- | --- | --- | --- | --- | --- | --- | --- |
| Probabilities | 0 - 0.140 | 0.141 - 0.25 | 0.251 - 0.37 | 0.371 - 0.46 | 0.461 - 0.54 | 0.541 - 0.62 | 0.621 - 0.72 | 0.721 - 0.81 | 0.811 - 0.91 | 0.911 - 1.0 |
| S to CVD ratio | 130 to 4 | 120 to 9 | 107 to 14 | 80 to 28 | 60 to 34 | 53 to 37 | 45 to 41 | 33 to 46 | 18 to 52 | 7 to 57 |
| CVD% | 2.9% | 7% | 11.6% | 25.9% | 36.2% | 41.1% | 47.7% | 54.4% | 74.3% | 89.1% |
| Given this data, the following base points will be used to fit a quadratic curve: |  |  |  |  |  |  |  |  |  |  |
| X | 0.07 | 0.19 | 0.31 | 0.415 | 0.5 | 0.58 | 0.67 | 0.765 | 0.86 | 0.96 |
| Y | 0.029 | 0.07 | 0.116 | 0.259 | 0.362 | 0.411 | 0.477 | 0.544 | 0.743 | 0.891 |

In addition the point 0/0 is added to the curve in order to achieve a sensible intercept.
This results in the following quadratic fit:
Quadratic fit with $y = ax + bx^2$ where $a = 0.3143$ and $b = 0.6226$ . which will be used to determine the percentage risk for a patient based on the predicted probability.

**Supplement Table S3.**
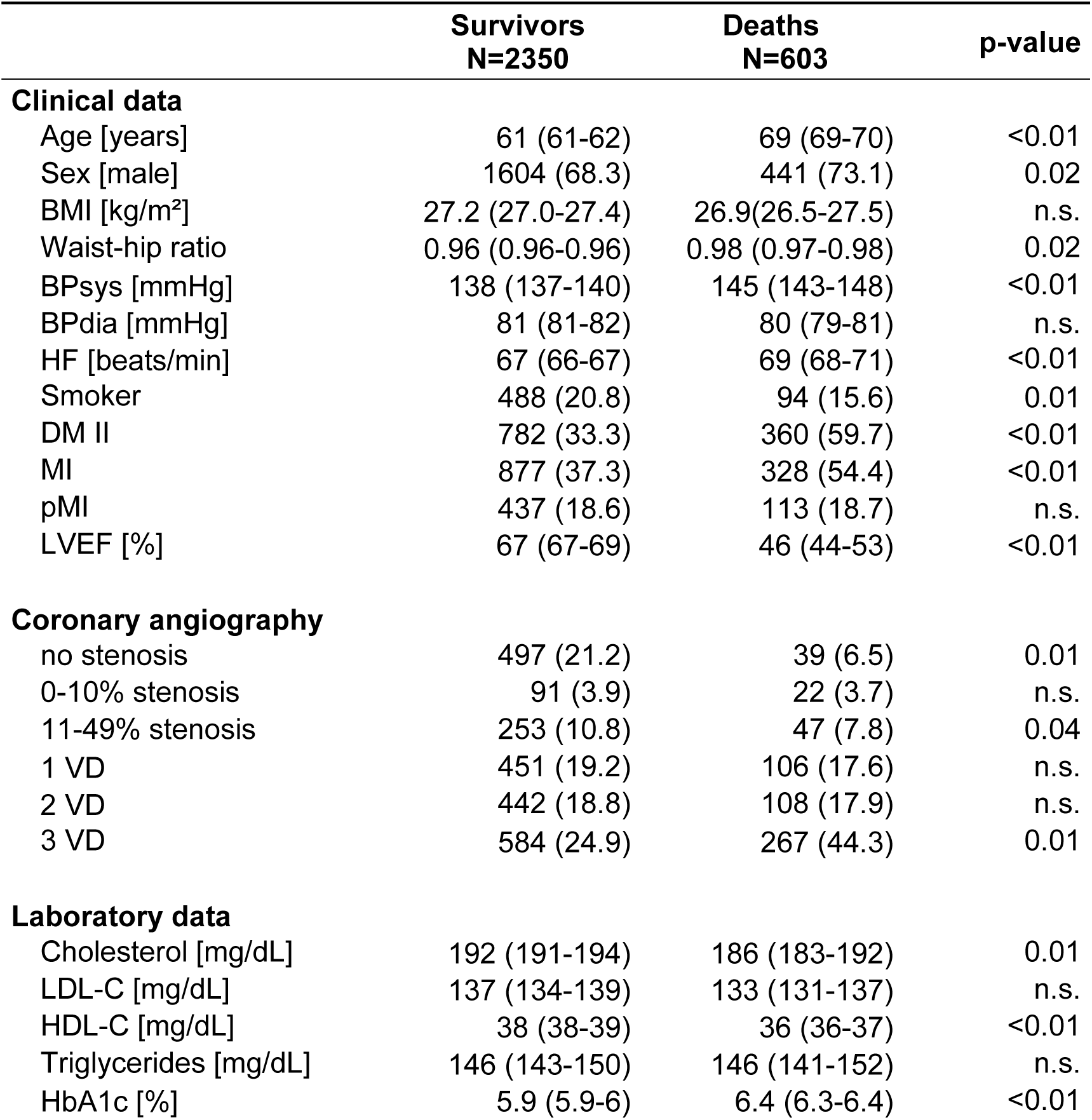
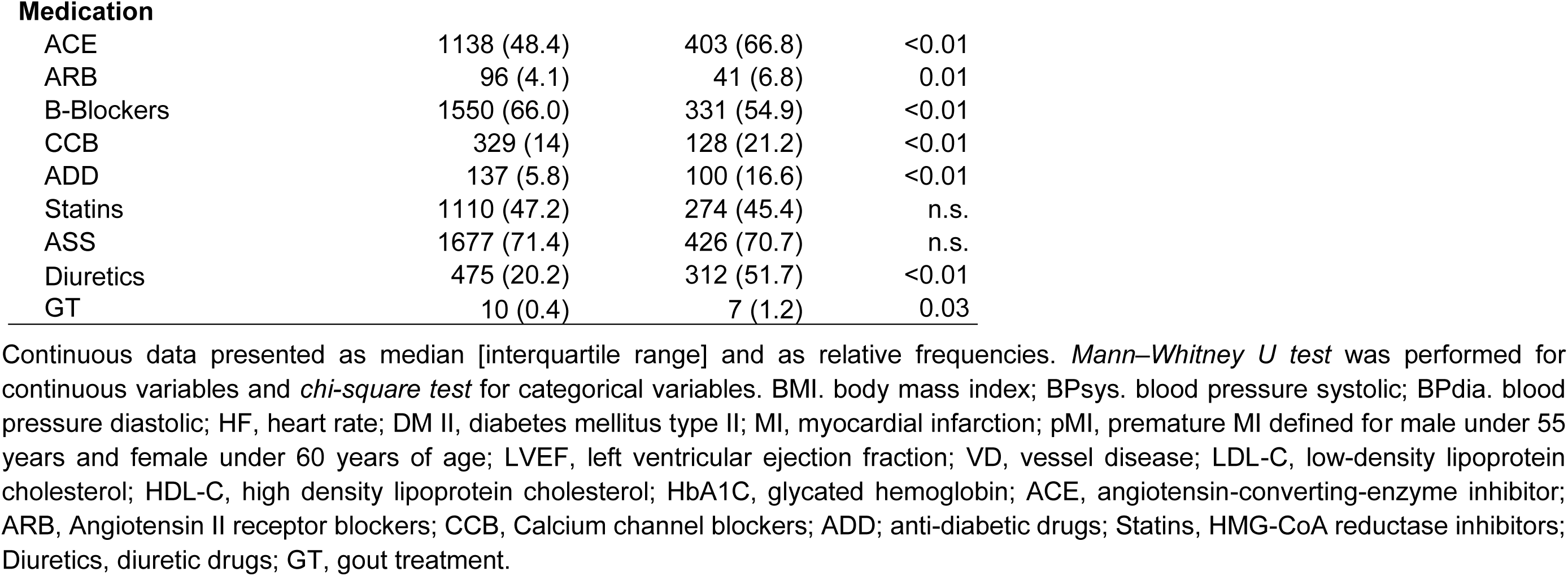
Baseline characteristics of overall study population.

|  | **Survivors**<br>**N=2350** | **Deaths**<br>**N=603** | **p-value** |
| --- | --- | --- | --- |
| **Clinical data** |  |  |  |
| Age [years] | 61 (61-62) | 69 (69-70) | <0.01 |
| Sex [male] | 1604 (68.3) | 441 (73.1) | 0.02 |
| BMI [kg/m<sup>2</sup>] | 27.2 (27.0-27.4) | 26.9(26.5-27.5) | n.s. |
| Waist-hip ratio | 0.96 (0.96-0.96) | 0.98 (0.97-0.98) | 0.02 |
| BPsys [mmHg] | 138 (137-140) | 145 (143-148) | <0.01 |
| BPdia [mmHg] | 81 (81-82) | 80 (79-81) | n.s. |
| HF [beats/min] | 67 (66-67) | 69 (68-71) | <0.01 |
| Smoker | 488 (20.8) | 94 (15.6) | 0.01 |
| DM II | 782 (33.3) | 360 (59.7) | <0.01 |
| MI | 877 (37.3) | 328 (54.4) | <0.01 |
| pMI | 437 (18.6) | 113 (18.7) | n.s. |
| LVEF [%] | 67 (67-69) | 46 (44-53) | <0.01 |
| **Coronary angiography** |  |  |  |
| no stenosis | 497 (21.2) | 39 (6.5) | 0.01 |
| 0-10% stenosis | 91 (3.9) | 22 (3.7) | n.s. |
| 11-49% stenosis | 253 (10.8) | 47 (7.8) | 0.04 |
| 1 VD | 451 (19.2) | 106 (17.6) | n.s. |
| 2 VD | 442 (18.8) | 108 (17.9) | n.s. |
| 3 VD | 584 (24.9) | 267 (44.3) | 0.01 |
| **Laboratory data** |  |  |  |
| Cholesterol [mg/dL] | 192 (191-194) | 186 (183-192) | 0.01 |
| LDL-C [mg/dL] | 137 (134-139) | 133 (131-137) | n.s. |
| HDL-C [mg/dL] | 38 (38-39) | 36 (36-37) | <0.01 |
| Triglycerides [mg/dL] | 146 (143-150) | 146 (141-152) | n.s. |
| HbA1c [%] | 5.9 (5.9-6) | 6.4 (6.3-6.4) | <0.01 |
| **Medication** |  |  |  |
| ACE | 1138 (48.4) | 403 (66.8) | <0.01 |
| ARB | 96 (4.1) | 41 (6.8) | 0.01 |
| B-Blockers | 1550 (66.0) | 331 (54.9) | <0.01 |
| CCB | 329 (14) | 128 (21.2) | <0.01 |
| ADD | 137 (5.8) | 100 (16.6) | <0.01 |
| Statins | 1110 (47.2) | 274 (45.4) | n.s. |
| ASS | 1677 (71.4) | 426 (70.7) | n.s. |
| Diuretics | 475 (20.2) | 312 (51.7) | <0.01 |
| GT | 10 (0.4) | 7 (1.2) | 0.03 |

Continuous data presented as median [interquartile range] and as relative frequencies. *Mann–Whitney U test* was performed for continuous variables and *chi-square test* for categorical variables. BMI, body mass index; BPsyst. blood pressure systolic; BPdia. blood pressure diastolic; HF, heart rate; DM II, diabetes mellitus type II; MI, myocardial infarction; pMI, premature MI defined for male under 55 years and female under 60 years of age; LVEF, left ventricular ejection fraction; VD, vessel disease; LDL-C, low-density lipoprotein cholesterol; HDL-C, high density lipoprotein cholesterol; HbA1C, glycated hemoglobin; ACE, angiotensin-converting-enzyme inhibitor; ARB, Angiotensin II receptor blockers; CCB, Calcium channel blockers; ADD; anti-diabetic drugs; Statins, HMG-CoA reductase inhibitors; Diuretics, diuretic drugs; GT, gout treatment.

**Supplement Table S4.**
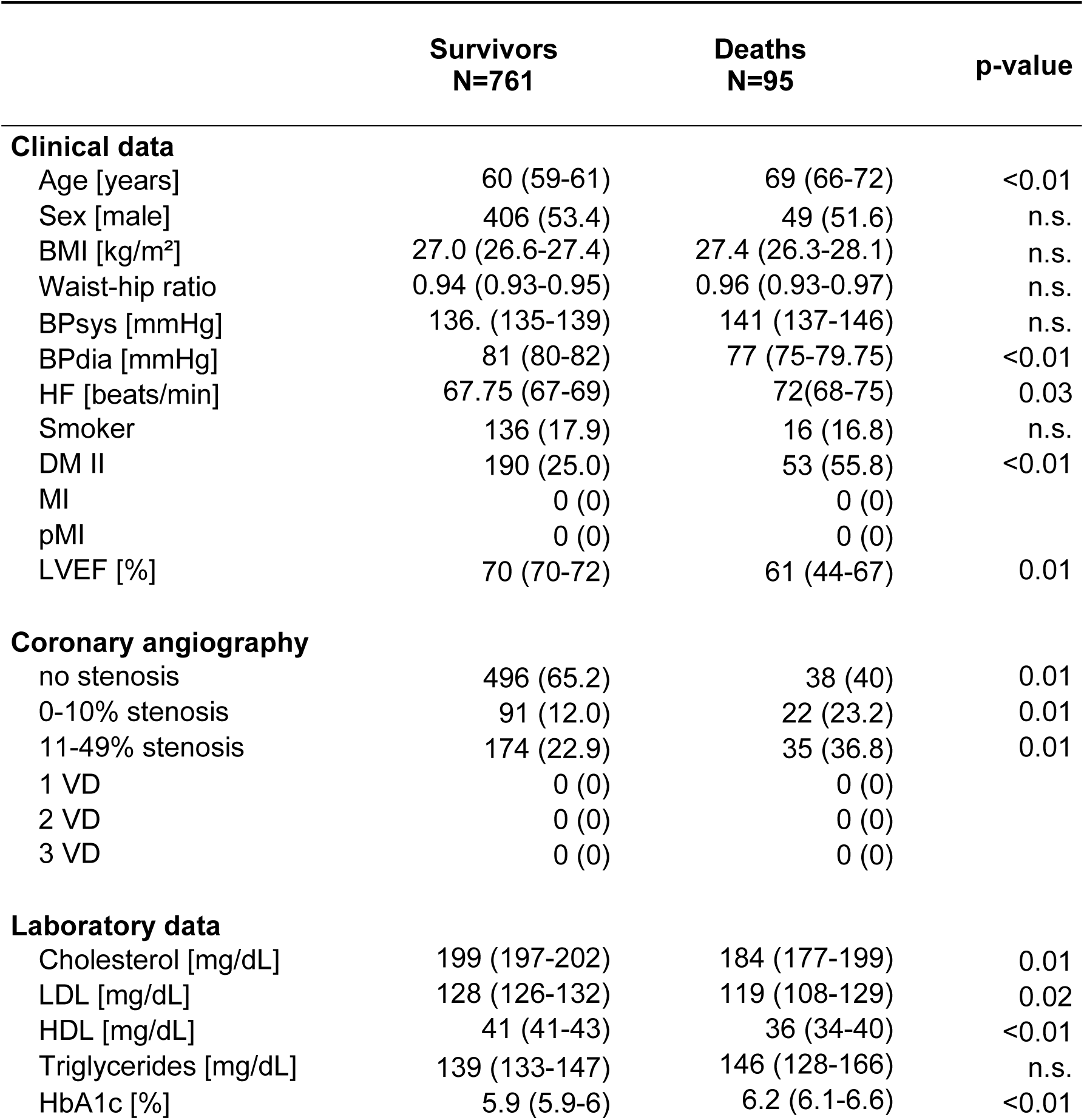
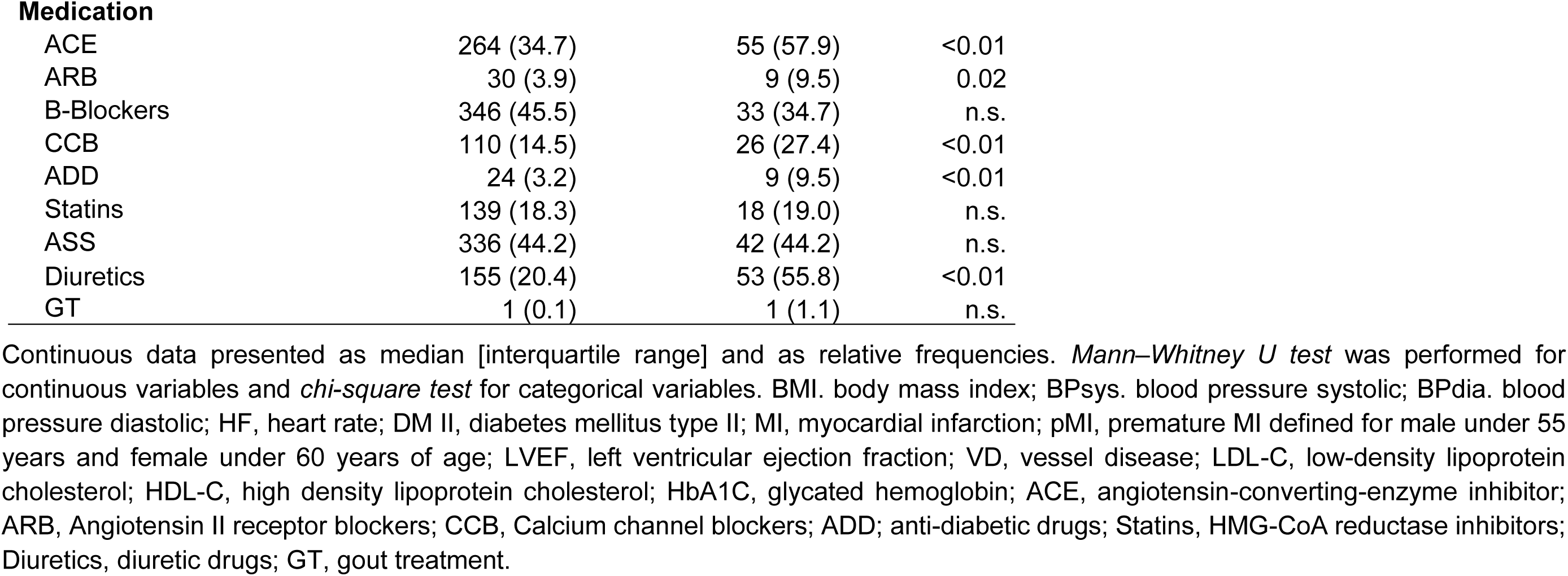
Baseline characteristics of patients without CAD.

|  | **Survivors**<br>**N=761** | **Deaths**<br>**N=95** | **p-value** |
| --- | --- | --- | --- |
| **Clinical data** |  |  |  |
| Age [years] | 60 (59-61) | 69 (66-72) | <0.01 |
| Sex [male] | 406 (53.4) | 49 (51.6) | n.s. |
| BMI [kg/m<sup>2</sup>] | 27.0 (26.6-27.4) | 27.4 (26.3-28.1) | n.s. |
| Waist-hip ratio | 0.94 (0.93-0.95) | 0.96 (0.93-0.97) | n.s. |
| BPsys [mmHg] | 136. (135-139) | 141 (137-146) | n.s. |
| BPdia [mmHg] | 81 (80-82) | 77 (75-79.75) | <0.01 |
| HF [beats/min] | 67.75 (67-69) | 72(68-75) | 0.03 |
| Smoker | 136 (17.9) | 16 (16.8) | n.s. |
| DM II | 190 (25.0) | 53 (55.8) | <0.01 |
| MI | 0 (0) | 0 (0) |  |
| pMI | 0 (0) | 0 (0) |  |
| LVEF [%] | 70 (70-72) | 61 (44-67) | 0.01 |
| **Coronary angiography** |  |  |  |
| no stenosis | 496 (65.2) | 38 (40) | 0.01 |
| 0-10% stenosis | 91 (12.0) | 22 (23.2) | 0.01 |
| 11-49% stenosis | 174 (22.9) | 35 (36.8) | 0.01 |
| 1 VD | 0 (0) | 0 (0) |  |
| 2 VD | 0 (0) | 0 (0) |  |
| 3 VD | 0 (0) | 0 (0) |  |
| **Laboratory data** |  |  |  |
| Cholesterol [mg/dL] | 199 (197-202) | 184 (177-199) | 0.01 |
| LDL [mg/dL] | 128 (126-132) | 119 (108-129) | 0.02 |
| HDL [mg/dL] | 41 (41-43) | 36 (34-40) | <0.01 |
| Triglycerides [mg/dL] | 139 (133-147) | 146 (128-166) | n.s. |
| HbA1c [%] | 5.9 (5.9-6) | 6.2 (6.1-6.6) | <0.01 |
| **Medication** |  |  |  |
| ACE | 264 (34.7) | 55 (57.9) | <0.01 |
| ARB | 30 (3.9) | 9 (9.5) | 0.02 |
| B-Blockers | 346 (45.5) | 33 (34.7) | n.s. |
| CCB | 110 (14.5) | 26 (27.4) | <0.01 |
| ADD | 24 (3.2) | 9 (9.5) | <0.01 |
| Statins | 139 (18.3) | 18 (19.0) | n.s. |
| ASS | 336 (44.2) | 42 (44.2) | n.s. |
| Diuretics | 155 (20.4) | 53 (55.8) | <0.01 |
| GT | 1 (0.1) | 1 (1.1) | n.s. |

Continuous data presented as median [interquartile range] and as relative frequencies. *Mann–Whitney U test* was performed for continuous variables and *chi-square test* for categorical variables. BMI, body mass index; BPsyst. blood pressure systolic; BPdia. blood pressure diastolic; HF, heart rate; DM II, diabetes mellitus type II; MI, myocardial infarction; pMI, premature MI defined for male under 55 years and female under 60 years of age; LVEF, left ventricular ejection fraction; VD, vessel disease; LDL-C, low-density lipoprotein cholesterol; HDL-C, high density lipoprotein cholesterol; HbA1C, glycated hemoglobin; ACE, angiotensin-converting-enzyme inhibitor; ARB, Angiotensin II receptor blockers; CCB, Calcium channel blockers; ADD; anti-diabetic drugs; Statins, HMG-CoA reductase inhibitors; Diuretics, diuretic drugs; GT, gout treatment.

**Supplement Table S5.**
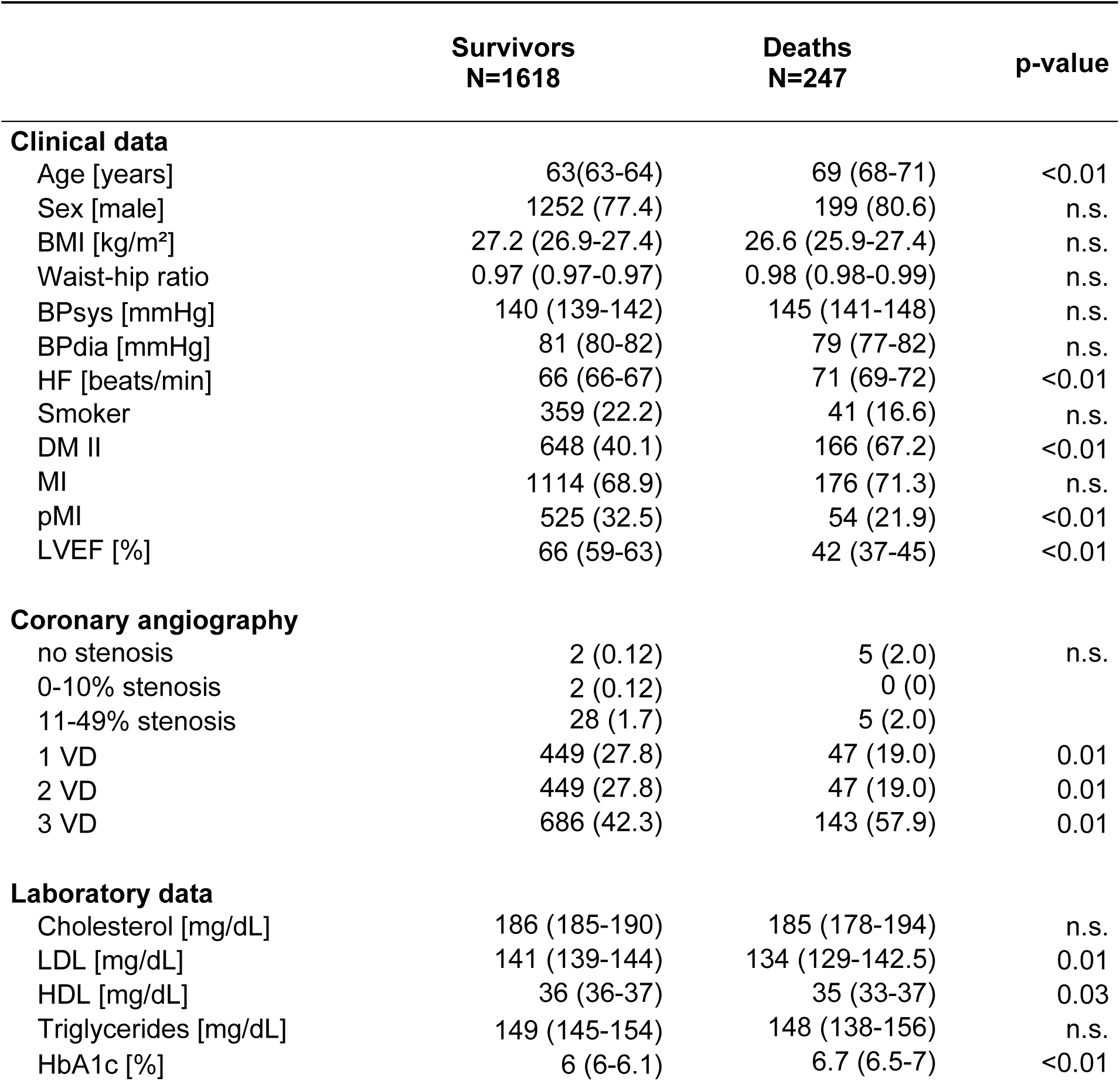
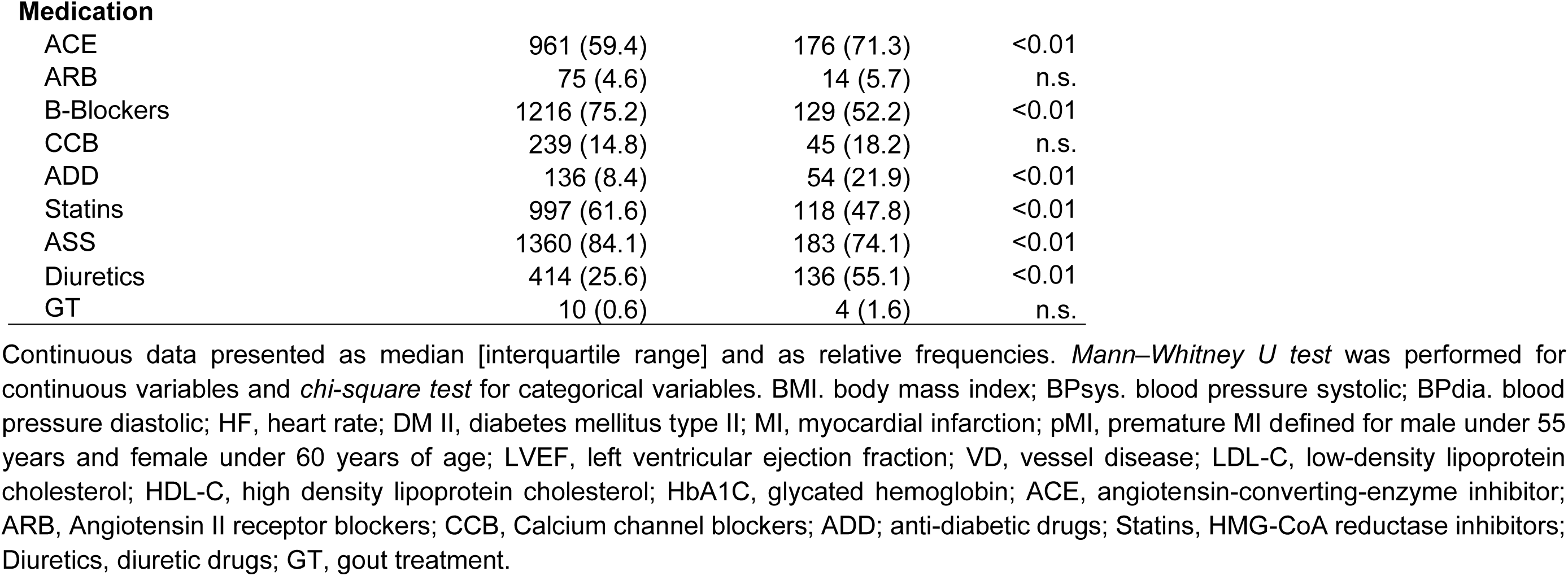
Baseline characteristics of the highCAD subpopulation.

|  | **Survivors**<br>**N=1618** | **Deaths**<br>**N=247** | **p-value** |
| --- | --- | --- | --- |
| **Clinical data** |  |  |  |
| Age [years] | 63(63-64) | 69 (68-71) | <0.01 |
| Sex [male] | 1252 (77.4) | 199 (80.6) | n.s. |
| BMI [kg/m<sup>2</sup>] | 27.2 (26.9-27.4) | 26.6 (25.9-27.4) | n.s. |
| Waist-hip ratio | 0.97 (0.97-0.97) | 0.98 (0.98-0.99) | n.s. |
| BPsys [mmHg] | 140 (139-142) | 145 (141-148) | n.s. |
| BPdia [mmHg] | 81 (80-82) | 79 (77-82) | n.s. |
| HF [beats/min] | 66 (66-67) | 71 (69-72) | <0.01 |
| Smoker | 359 (22.2) | 41 (16.6) | n.s. |
| DM II | 648 (40.1) | 166 (67.2) | <0.01 |
| MI | 1114 (68.9) | 176 (71.3) | n.s. |
| pMI | 525 (32.5) | 54 (21.9) | <0.01 |
| LVEF [%] | 66 (59-63) | 42 (37-45) | <0.01 |
| **Coronary angiography** |  |  |  |
| no stenosis | 2 (0.12) | 5 (2.0) | n.s. |
| 0-10% stenosis | 2 (0.12) | 0 (0) |  |
| 11-49% stenosis | 28 (1.7) | 5 (2.0) |  |
| 1 VD | 449 (27.8) | 47 (19.0) | 0.01 |
| 2 VD | 449 (27.8) | 47 (19.0) | 0.01 |
| 3 VD | 686 (42.3) | 143 (57.9) | 0.01 |
| **Laboratory data** |  |  |  |
| Cholesterol [mg/dL] | 186 (185-190) | 185 (178-194) | n.s. |
| LDL [mg/dL] | 141 (139-144) | 134 (129-142.5) | 0.01 |
| HDL [mg/dL] | 36 (36-37) | 35 (33-37) | 0.03 |
| Triglycerides [mg/dL] | 149 (145-154) | 148 (138-156) | n.s. |
| HbA1c [%] | 6 (6-6.1) | 6.7 (6.5-7) | <0.01 |
| **Medication** |  |  |  |
| ACE | 961 (59.4) | 176 (71.3) | <0.01 |
| ARB | 75 (4.6) | 14 (5.7) | n.s. |
| B-Blockers | 1216 (75.2) | 129 (52.2) | <0.01 |
| CCB | 239 (14.8) | 45 (18.2) | n.s. |
| ADD | 136 (8.4) | 54 (21.9) | <0.01 |
| Statins | 997 (61.6) | 118 (47.8) | <0.01 |
| ASS | 1360 (84.1) | 183 (74.1) | <0.01 |
| Diuretics | 414 (25.6) | 136 (55.1) | <0.01 |
| GT | 10 (0.6) | 4 (1.6) | n.s. |

Continuous data presented as median [interquartile range] and as relative frequencies. *Mann–Whitney U test* was performed for continuous variables and *chi-square test* for categorical variables. BMI, body mass index; BPsyst. blood pressure systolic; BPdia. blood pressure diastolic; HF, heart rate; DM II, diabetes mellitus type II; MI, myocardial infarction; pMI, premature MI defined for male under 55 years and female under 60 years of age; LVEF, left ventricular ejection fraction; VD, vessel disease; LDL-C, low-density lipoprotein cholesterol; HDL-C, high density lipoprotein cholesterol; HbA1C, glycated hemoglobin; ACE, angiotensin-converting-enzyme inhibitor; ARB, Angiotensin II receptor blockers; CCB, Calcium channel blockers; ADD; anti-diabetic drugs; Statins, HMG-CoA reductase inhibitors; Diuretics, diuretic drugs; GT, gout treatment.

